# Translating Subphenotypes of Newly Diagnosed Type 2 Diabetes from Cohort Studies to Electronic Health Records in the United States

**DOI:** 10.1101/2024.10.08.24315128

**Authors:** Zhongyu Li, Star Liu, Joyce C. Ho, K.M. Venkat Narayan, Mohammed K. Ali, Jithin Sam Varghese

## Abstract

Novel subphenotypes of type 2 diabetes mellitus (T2DM) are associated with differences in response to treatment and risk of complications. The most widely replicated approach identified four subphenotypes (severe insulin-deficient diabetes [SIDD], severe insulin-resistant diabetes [SIRD], mild obesity-related diabetes [MOD], and mild age-related diabetes [MARD]). However, the widespread clinical application of this model is hindered by the limited availability of fasting insulin and glucose measurements in routine clinical settings. To address this, we pooled data of adults (≥18 years) with newly diagnosed T2DM from six cohort studies (n = 3,377) to perform de novo clustering and developed classification algorithms for each of the four subphenotypes using nine variables routinely collected in electronic health records (EHRs). After operationalizing the classification algorithms on the Epic Cosmos Research Platform, we identified that among the 727,076 newly diagnosed diabetes cases, 21.6% were classified as SIDD, 23.8% as MOD, and 40.9% as MARD. Individuals classified as SIDD were more likely to receive insulin and incretin mimetics treatment and had higher risks for microvascular complications (retinopathy, neuropathy, nephropathy). Our findings underscore the heterogeneity in newly diagnosed T2DM and validated T2DM subphenotypes in routine EHR systems. This offers possibilities for the subsequent development of treatment strategies tailored to subphenotypes.

## INTRODUCTION

Type 2 Diabetes Mellitus (T2DM) affects over 500 million people worldwide, including over 34 million adults in the United States (US).^1,2^ Recent studies using unsupervised machine learning identified subphenotypes of T2DM with differences in pathophysiology, response to medication, and risk of microvascular complications.^3,4,5^ Although several approaches were employed for subphenotyping of T2DM, the most widely replicated study from the Swedish All New Diabetics in Scandia (ANDIS) cohort identified four subphenotypes, namely Severe Insulin Deficient Diabetes (SIDD), Severe Insulin Resistant Diabetes (SIRD), Mild Obesity-related Diabetes (MOD) and Mild Age-related Diabetes (MARD).^6–8^ To date, these subphenotypes have not yet been translated into health gains from better risk stratification through precision medicine.^9,10^

At least two gaps exist in clinical validation and subsequent translation to real-world clinical practice. First, not all variables used for subphenotyping are routinely collected during T2DM diagnosis at clinic visits.^11^ A majority of studies that identified the ANDIS subphenotypes were conducted in observational cohort studies,^12^ where homeostatic indices for insulin secretion and resistance that used fasting levels of c-peptide or insulin are collected.^12^ As a result, it remains unknown whether these subphenotypes can be replicated using variables available in electronic health records (EHRs). Second, existing data-driven classification from cohort studies in European countries may not be generalizable across geographies and ethnic groups, including diverse populations of the US and Asia. For instance, studies in Asian cohorts have found differences from Europeans in their relative composition of phenotypes (higher proportions of SIDD), but also identified additional phenotypes such as Combined Insulin Deficient and Resistant Diabetes.^7,13,14^ Additionally, there are variations in subphenotype characteristics even among geographically and ethnically similar populations in Europe.^6,15,16^ Therefore, there is a critical need to identify de-novo clusters in ethnically and socio-demographically diverse source populations from the US and subsequently enable validation in clinical settings.^17^

To address these knowledge gaps in subphenotyping newly diagnosed T2DM and study their epidemiology in EHRs, we implemented a two-step approach using pooled data from six diverse cohort studies and a large integrated EHR database in the US (**Figure 1**). First, we derived novel subphenotypes of newly diagnosed T2DM in cohort studies and developed classification algorithms for each subphenotype using variables routinely collected in EHRs. Second, we applied the classification algorithms for subphenotypes in EHRs and studied the time to the prescription of pharmacological treatments and the incidence of microvascular complications.

**Figure 1.**
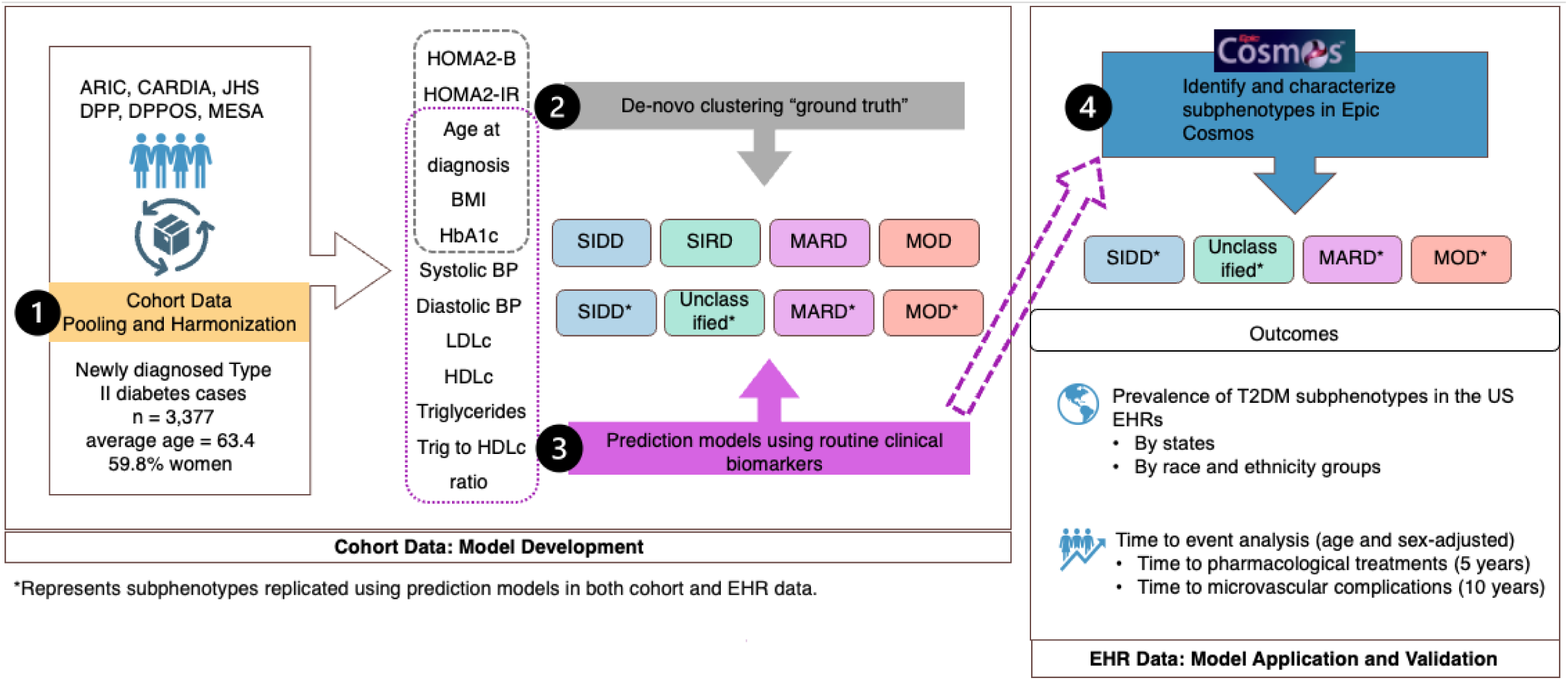
Study design for translating subphenotypes from cohort studies to electronic health records.

**Figure 2.**
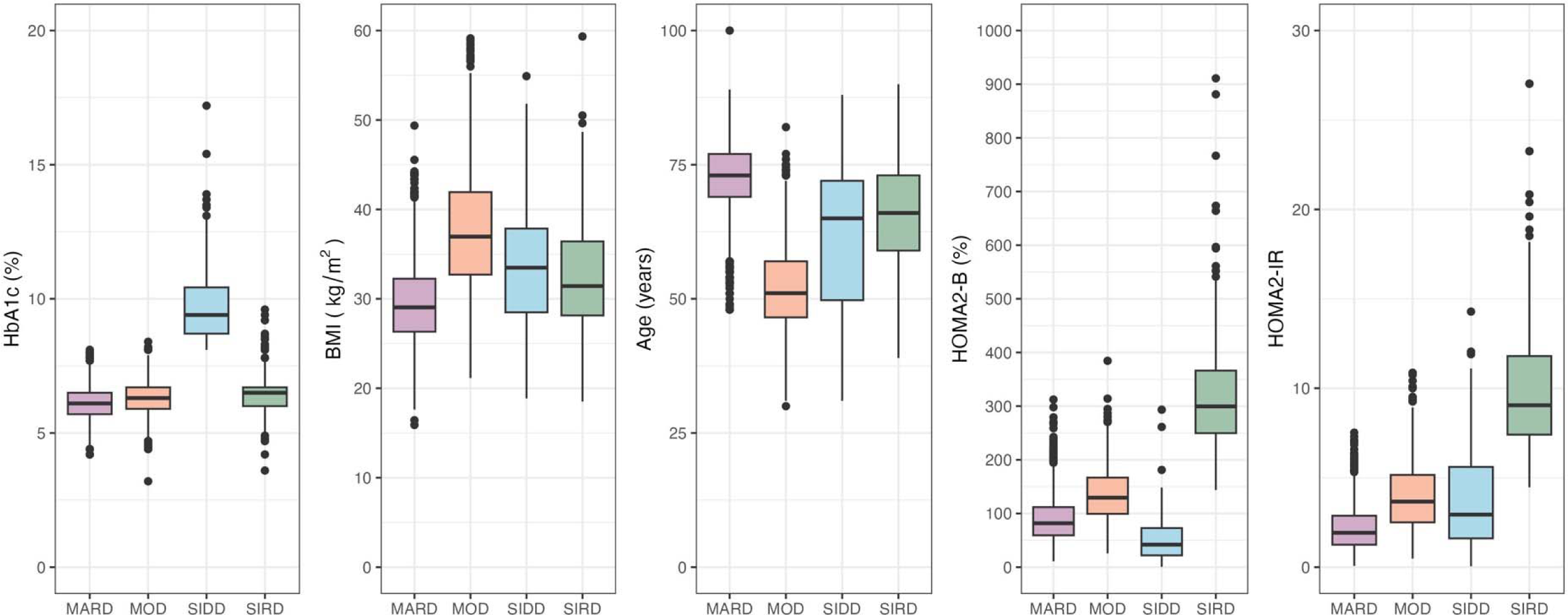
Distribution of key variables by subphenotypes of newly diagnosed type 2 diabetes in pooled cohort studies. Distribution of HbA1c (%), BMI (kg/m^2^), age (years), HOMA2-B (%), HOMA2-IR for de-novo clusters at diagnosis. K-means clustering was not done separately for males and females based on findings from ANDIS study that suggested similarities in cluster centroids by sex.

## RESULTS

### Participant Characteristics in the Pooled Cohorts

We pooled data of newly diagnosed T2DM from six US cohort studies: the Atherosclerosis Risk in Communities (ARIC) Study, Coronary Artery Risk Development in Young Adults Study (CARDIA), Diabetes Prevention Program (DPP) and Diabetes Prevention Program Outcomes Study (DPPOS), Jackson Heart Study (JHS), and the Multi-Ethnic Study of Atherosclerosis (MESA). These cohorts enrolled socio-demographically and geographically diverse populations with varying clinical profiles (**Supplementary Table 1**), enabling us to capture a broad spectrum of diabetes presentations.

Among these studies, the DPPOS is the observational follow-up to the randomized controlled trial, DPP. We excluded participants from the intervention arms of the DPP for the main analysis. This exclusion was done to minimize the potential effects of exposure to diabetes interventions, such as lifestyle modifications or medications, on the biomarkers used in our classification algorithms. Besides, to ensure the independence of the JHS cohort and avoid double counting, we excluded participants who were also recruited in the ARIC study from the JHS cohort. All studies used internally standardized protocols to measure anthropometry and biomarkers (**Supplementary Table 2**), routinely assessed dysglycemia in their participants, and had a low risk of left censoring of newly diagnosed T2DM.

After harmonizing definitions across six U.S.-based cohort studies (Supplementary Table 3), we identified 7,623 participants with newly diagnosed T2DM. We then excluded individuals missing key biomarkers, including body mass index (BMI), age at diagnosis, and glycated hemoglobin (HbA1c), as well as those with implausible homeostatic assessments at diagnosis (n = 13). This resulted in an analytic sample of 3,377 newly diagnosed T2DM cases for developing the classification algorithm in the pooled cohort dataset (**Supplementary Figure 1**).

Descriptive characteristics of the pooled cohort sample and each cohort study are presented in **Table 1** and **Supplementary Table 4**. The average age of participants at diagnosis of T2DM was 63.4 years (SD: 12.4), with younger ages at diagnosis in CARDIA (48.8 years [SD: 4.9]) and older age at diagnosis in ARIC (75.3 years [SD: 5.1]). The overall pooled cohort were 59.8% female, 49.9% White, 35.5% Black and 14.6% from other racial groups with average HbA1c of 6.3% (IQR: 5.8-6.7%) and BMI of 33.2 kg/m^2^ (SD: 7.0). ARIC was predominantly White (72.1%) while CARDIA and JHS had higher proportions of Black participants (70.6% and 100% respectively) (**Supplementary Table 4**). The pooled cohort had high median values of insulin resistance and beta cell function, defined using homeostatic model assessment indices of HOMA2-IR (2.8 [interquartile range: 1.7-4.7]) and HOMA2-B% (108% [IQR: 73.3-156.4]). There was variability across the six cohort studies for all metabolic biomarkers (range of median values; systolic blood pressure [SBP]: 106.23 to 129.6 mmHg, diastolic blood pressure [DBP]: 66.8 to 99.5 mmHg, LDL cholesterol: 97.3 to 121.5 mg/dL, HDL cholesterol: 42.1 to 49.5 mg/dL, triglycerides: 128.8 to 175.3 mg/dL, triglyceride-to-HDL ratio: 2.1 to 3.6).

**Table 1.**
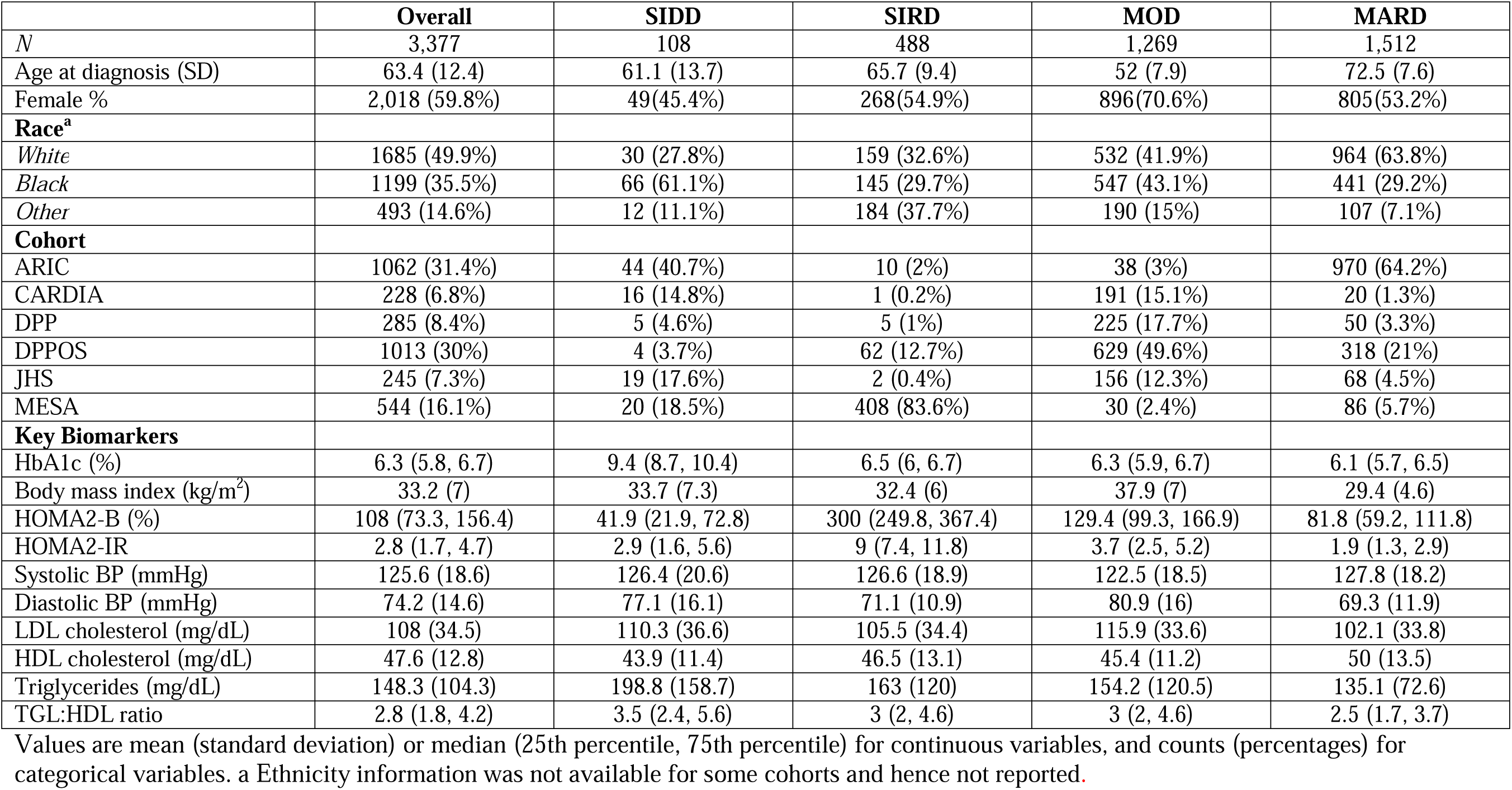
Descriptive characteristics of newly diagnosed T2DM subphenotypes in pooled cohort studies.

|  | **Overall** | **SIDD** | **SIRD** | **MOD** | **MARD** |
| --- | --- | --- | --- | --- | --- |
| *N* | 3,377 | 108 | 488 | 1,269 | 1,512 |
| Age at diagnosis (SD) | 63.4 (12.4) | 61.1 (13.7) | 65.7 (9.4) | 52 (7.9) | 72.5 (7.6) |
| Female % | 2,018 (59.8%) | 49(45.4%) | 268(54.9%) | 896(70.6%) | 805(53.2%) |
| **Race<sup>a</sup>** |  |  |  |  |  |
| *White* | 1685 (49.9%) | 30 (27.8%) | 159 (32.6%) | 532 (41.9%) | 964 (63.8%) |
| *Black* | 1199 (35.5%) | 66 (61.1%) | 145 (29.7%) | 547 (43.1%) | 441 (29.2%) |
| *Other* | 493 (14.6%) | 12 (11.1%) | 184 (37.7%) | 190 (15%) | 107 (7.1%) |
| **Cohort** |  |  |  |  |  |
| ARIC | 1062 (31.4%) | 44 (40.7%) | 10 (2%) | 38 (3%) | 970 (64.2%) |
| CARDIA | 228 (6.8%) | 16 (14.8%) | 1 (0.2%) | 191 (15.1%) | 20 (1.3%) |
| DPP | 285 (8.4%) | 5 (4.6%) | 5 (1%) | 225 (17.7%) | 50 (3.3%) |
| DPPOS | 1013 (30%) | 4 (3.7%) | 62 (12.7%) | 629 (49.6%) | 318 (21%) |
| JHS | 245 (7.3%) | 19 (17.6%) | 2 (0.4%) | 156 (12.3%) | 68 (4.5%) |
| MESA | 544 (16.1%) | 20 (18.5%) | 408 (83.6%) | 30 (2.4%) | 86 (5.7%) |
| **Key Biomarkers** |  |  |  |  |  |
| HbA1c (%) | 6.3 (5.8, 6.7) | 9.4 (8.7, 10.4) | 6.5 (6, 6.7) | 6.3 (5.9, 6.7) | 6.1 (5.7, 6.5) |
| Body mass index (kg/m<sup>2</sup>) | 33.2 (7) | 33.7 (7.3) | 32.4 (6) | 37.9 (7) | 29.4 (4.6) |
| HOMA2-B (%) | 108 (73.3, 156.4) | 41.9 (21.9, 72.8) | 300 (249.8, 367.4) | 129.4 (99.3, 166.9) | 81.8 (59.2, 111.8) |
| HOMA2-IR | 2.8 (1.7, 4.7) | 2.9 (1.6, 5.6) | 9 (7.4, 11.8) | 3.7 (2.5, 5.2) | 1.9 (1.3, 2.9) |
| Systolic BP (mmHg) | 125.6 (18.6) | 126.4 (20.6) | 126.6 (18.9) | 122.5 (18.5) | 127.8 (18.2) |
| Diastolic BP (mmHg) | 74.2 (14.6) | 77.1 (16.1) | 71.1 (10.9) | 80.9 (16) | 69.3 (11.9) |
| LDL cholesterol (mg/dL) | 108 (34.5) | 110.3 (36.6) | 105.5 (34.4) | 115.9 (33.6) | 102.1 (33.8) |
| HDL cholesterol (mg/dL) | 47.6 (12.8) | 43.9 (11.4) | 46.5 (13.1) | 45.4 (11.2) | 50 (13.5) |
| Triglycerides (mg/dL) | 148.3 (104.3) | 198.8 (158.7) | 163 (120) | 154.2 (120.5) | 135.1 (72.6) |
| TGL:HDL ratio | 2.8 (1.8, 4.2) | 3.5 (2.4, 5.6) | 3 (2, 4.6) | 3 (2, 4.6) | 2.5 (1.7, 3.7) |

Values are mean (standard deviation) or median (25th percentile, 75th percentile) for continuous variables, and counts (percentages) for categorical variables. a Ethnicity information was not available for some cohorts and hence not reported.

### De-novo clustering of Subphenotypes of Incident Diabetes in the Pooled Cohorts

Consistent with the other studies of subphenotypes of newly diagnosed T2DM, we conducted a de-novo hierarchical clustering and k-means clustering utilizing five key variables: age at diagnosis, BMI, HbA1c (%), HOMA2 %B, and HOMA2 IR. Both approaches identified an optimal solution of four clusters (**Supplementary Figure 2**). The clusters identified through k-means were subsequently labeled according to their similarity in the distribution of the five clustering variables (**Figure 1**) to those subphenotypes described in the original ANDIS study: Mild Obesity-Related Diabetes (MOD), Mild Age-Related Diabetes (MARD), Severe Insulin-Deficient Diabetes (SIDD), and Severe Insulin-Resistant Diabetes (SIRD). A sensitivity analysis showed that individuals were predominantly classified into the same four subphenotypes when using cluster centroids from the ANDIS study (**Supplementary Table 5**).

In the pooled cohort sample, MARD was the most common subphenotype, representing 44.5% of those with newly diagnosed T2DM, followed by MARD (37.5%), SIRD (14.5%) and SIDD (3.2%). Cases identified as MARD were older (72.5 years [SD: 7.6]) with lower BMI (29.4 kg/m^2^ [SD: 4.6]) and HOMA2-IR (1.9 [IQR: 1.3-2.9]). MOD was characterized by younger age (52 years [SD: 7.9]), higher BMI (37.9 kg/m² [SD: 7.0]) and a high HOMA2-IR (3.7 [IQR: 2.5-5.2]) (**Table 1**). The majority of newly diagnosed T2DM in CARDIA (83.8%), DPP (79.8%), DPPOS (62.1%), and JHS (63.7%) were identified as MOD (**Supplementary Table 4**).

Nearly all newly diagnosed cases in ARIC (91.3%) were identified as MARD. The SIDD and SIRD subphenotypes had worse cardiometabolic profiles, compared to MOD and MARD. SIRD was characterized by high BMI (32.4 kg/m^2^ [SD: 5.8]), HOMA2-B% (303 [IQR: 250.6-374.8]) and HOMA2-IR (9.2 [IQR: 7.4-11.8]). SIRD also had a later age at diagnosis (65.7 years [SD: 9.4]) than MOD and SIDD but earlier than MARD. SIDD was marked by early age at onset (61.1 years [SD: 13.7]), high HbA1c (9.4% [IQR: 8.7-10.4]) and poor beta-cell function (HOMA2-B%: 41.9 [IQR: 21.9-72.8]).

To evaluate the robustness of the clustering algorithms to pooling cohort studies, we first examined whether the inclusion of any single large cohort meaningfully influenced the results. We calculated the Adjusted Rand Index (ARI) and Cohen’s κ between the original classification from the pooled cohort studies and those obtained using a leave-one-cohort-out approach. Both the ARI and Cohen’s κ values suggest a high level of stability and concordance (ARI ≥ 0.96, κ ≥ 0.98) in diabetes subphenotype clustering when excluding CARDIA, DPP, DPPOS and JHS, and moderate stability when excluding ARIC (ARI = 0.49, κ = 0.69) and MESA (ARI = 0.63, κ = 0.68) (**Supplementary Table 5**). Second, to examine if the imputation of missing clustering variables (i.e., blood pressure, LDL cholesterol, HDL cholesterol, triglycerides) using k-nearest neighbors imputation influenced the results, we conducted a de-novo clustering using complete cases (**Supplementary Table 7**). Subphenotypes identified using the pooled analytic sample (N= 3,377) and the complete case sample ( N = 2,775) displayed high concordance (**Supplementary Table 8**).

### Classification of Subphenotypes Using Routine Clinical Variables from the EHR

Replication studies of the four subphenotypes are usually conducted using HOMA2 indices, which are based on fasting insulin or C-peptide levels. However, these measurements are not commonly collected in routine clinical care (**Supplementary Table 9**), thereby limiting the validation and translation of these novel subphenotypes to clinical practice. To address this challenge, we explored alternative methods to classify T2DM subgroups using only routinely available clinical variables. First, we conducted unsupervised k-means clustering of the analytic sample using biomarkers available in EHRs, namely age of diagnosis, HbA1c, BMI, blood pressure, HDL, LDL, triglycerides, and triglycerides to HDL ratio (a proxy for insulin resistance).^18^ However, this approach yielded clusters that exhibited low concordance with the four ANDIS subphenotypes identified in our de-novo clustering analysis (**Supplementary Table 10**).

Next, we sought to refine the classification process by developing and validating four One-vs-All logistic regression models using the same routinely available clinical variables. This supervised learning approach allowed us to model an individual’s probability of membership in each subphenotype. Each model was fitted on a 70% training subset (n = 2,363) of the pooled cohort dataset and subsequently validated on a 30% held-out test dataset (n = 1,014). To determine the optimal cutoff to predict subphenotype membership, we employed a fivefold cross-validation approach and selected the predicted probability threshold that maximizes the F1 score. These thresholds were validated on the held-out test dataset to evaluate discrimination for each subphenotype. Key indicators of discrimination for each model in the training dataset and test dataset, i.e. sensitivity, specificity, area under the receiver operating characteristic curve (AUC), positive predictive value (PPV), negative predictive value (NPV), and F1 score, are presented in **Table 2**. Coefficients and standard errors for the logistic regression models are presented in **Supplementary Table 11.**

**Table 2.** Descriptive characteristics of newly detected type 2 diabetes by subphenotype in Epic Cosmos.

|  | **Overall** | **SIDD** | **MARD** | **MOD** | **Unclassified** |
| --- | --- | --- | --- | --- | --- |
|  | *727,076* | *156,951* | *297,568* | *172,922* | *99,635* |
| Age at detection of SUPREME-DM | 64.4 (13.3) | 59.9 (13.5) | 74.8(7.9) | 51.9 (9.4) | 62.9 (5.7) |
| Female | 52% | 50% | 50% | 58% | 49% |
| **Race & Ethnicity** |  |  |  |  |  |
| *NH White* | 68% | 63% | 73% | 61% | 69% |
| *NH Black* | 17% | 20% | 13% | 21% | 15% |
| *Hispanic* | 7% | 9.3% | 4.8% | 8.9% | 6.8% |
| *NH Other* | 8.6% | 8.0% | 8.8% | 8.5% | 8.8% |
| Social Vulnerability Index (0: Low, 100: High vulnerability) | 60.4 (33.5, 82.1) | 64.1 (37.0, 84.6) | 57.4(31.2, 79.7) | 62.7(35.5, 83.8) | 59.6 (32.8, 81.4) |
| **Insurance** |  |  |  |  |  |
| *Medicare* | 40% | 31% | 62% | 15% | 32% |
| *Medicaid* | 14% | 16% | 18% | 18% | 13% |
| **Key Biomarkers** |  |  |  |  |  |
| HbA1c (%) | 7.0 (6.6, 7.9) | 9.5 (8.7, 11.0) | 6.7 (6.5, 7.0) | 6.8 (6.5, 7.2) | 7.1 (6.8, 7.6) |
| Body mass index | 33.2 (6.8) | 34.0 (6.9) | 29.5 (5.1) | 38.8 (6.1) | 33.8 (5.3) |
| Systolic BP | 131.2 (15.5) | 132.5 (16.3) | 131.1(15.7) | 130.9 (14.8) | 129.9 (15.0) |
| Diastolic BP | 74.7 (10.2) | 76.2 (10.4) | 71.4 (9.5) | 78.7 (9.7) | 74.9 (9.3) |
| LDL cholesterol | 89.0 (37.8) | 96.2 (41.7) | 81.2 (34.8) | 97.5 (37.2) | 84.5 (35.5) |
| HDL cholesterol | 44.9 (14.6) | 42.9 (15.1) | 47.8 (15.1) | 43.7 (13.0) | 42.3 (13.9) |
| Triglycerides | 165.0 (92.4) | 185.9(106.2) | 141.6 (73.1) | 173.3 (93.5) | 183.9 (101.8) |
| TGL:HDL ratio | 3.3 (2.1, 5.3) | 3.9 (2.4, 6.4) | 2.8 (1.8, 4.2) | 3.6 (2.3, 5.5) | 3.9 (2.5, 6.4) |

Values are mean (standard deviation) or median (25<sup>th</sup> percentile, 75<sup>th</sup> percentile) for continuous variables, and percentages for categorical variables. Number of observations where biomarkers are missing is presented in **Supplementary Table 8**.

The models demonstrated strong overall performance, with minimal loss of discrimination between the training and test datasets (**Supplementary Table 12**). The optimal predicted probability thresholds for best classification based on the five-fold cross-validation were 0.16, 0.38, and 0.32, respectively, for SIDD, MOD, and MARD. In particular, the model for classifying SIDD exhibited high discrimination: AUC (0.99, 95% CI = 0.99, 1.00), sensitivity (0.94), specificity (0.99), and F1 score (0.90). Similarly, the models for MOD and MARD showed high sensitivity of 0.93 and 0.95, with F1 scores of 0.88 and 0.87, respectively, though they had lower specificity (MOD: 0.89, MARD: 0.81) compared to the SIDD model. The classification model for SIRD showed lower performance across all metrics, with an AUC of 0.62 (95% CI = 0.58, 0.67) and F1 score of 0.28. If an individual’s membership probability in a subphenotype was higher than the optimal threshold, we classified them sequentially as SIDD, MARD, and MOD based on the known risk of complications. Given lower discrimination indices for the SIRD model in reliably distinguishing this subphenotype from others, individuals not classified into any of the other three groups were categorized as “Unclassified”. Alternative classification approaches for subphenotypes (multinomial regression, random forests) did not improve discrimination (**Supplementary Table 13**).

### Epidemiology of Diabetes Subphenotype

Based on the high overall discrimination of SIDD, MARD and MOD in our pooled cohort analysis and previously reported associations of SIDD with a higher risk of diabetic complications, we subsequently applied the classification models to newly diagnosed T2DM cases (n = 727,076) identified in the Epic Cosmos Research Platform. This approach enabled us to characterize the epidemiology of subphenotypes in a diverse, real-world patient population, providing critical insights into its demographic, clinical, and treatment profiles. Newly diagnosed T2DM between January 2012 and December 2023 were identified using the SUPREME-DM computable phenotype. The newly diagnosed cases were on average 64.4 years (SD = 13.3), 52% female, 68% Non-Hispanic [NH] White (68%) 17% NH Black, 7% Hispanic and 8.6% NH Other, with an average BMI of 33.2 kg/m^2^ (SD: 6.8) and median HbA1c of 7.0% (IQR: 6.6-7.9). Median values of BMI, HbA1c and other biomarkers were similar to those observed in the pooled cohort data.

Applying logistic regression to predict subphenotype membership in Epic Cosmos identified 156,951 individuals (21.6%) as SIDD, 172,922 (23.8%) as MOD, and 297,568 (40.9%) as MARD. The average age at diagnosis was least for MOD (51.9 years [SD: 9.4]), followed by SIDD (59.8 years [SD: 13.5]), the unclassified group (62.9 years [SD: 5.7]), and MARD (74.8 years [SD: 7.9]) (**Table 2**). MOD had the highest average BMI (38.7 kg/m² [SD: 6.1]), while MARD had the lowest average BMI (29.4 kg/m² [SD: 4.0]). SIDD had the highest average HbA1c at diagnosis (9.5% [IQR: 8.7-11.0]), meaningfully higher than other subphenotypes, which ranged from 6.7% (MARD) to 7.1% (Unclassified). The unclassified group had a cardiometabolic profile resembling that of the MOD group, particularly in terms of BMI, blood pressure, and triglyceride to HDL cholesterol ratio. The social vulnerability index was higher for SIDD (64.1 [IQR: 37.0, 84.6]) and MOD (62.8 [IQR: 35.6, 83.8]) compared to MARD (57.3 [IQR: 31.1, 79.6])

The distribution of SIDD, MOD, and MARD among newly diagnosed type 2 diabetes cases across the US shows regional variations (**Figure 3**). For instance, the District of Columbia (27.8%) and South Carolina (26.1%) had the highest proportions of cases classified as SIDD, while Idaho (18.7%) and Kansas (18.9%) had the lowest proportions (**Figure 3, Panel A**). MARD is widespread across much of the U.S., with high prevalence in the Midwest (South Dakota: 49.4%), and the Northeast (Rhode Island: 46.6%) (**Figure 3, Panel B**). Utah showed the highest prevalence of MOD (29.2%), followed by Colorado (28.7%) and Alaska (27.4%) (**Figure 3, Panel C**).

**Figure 3.**
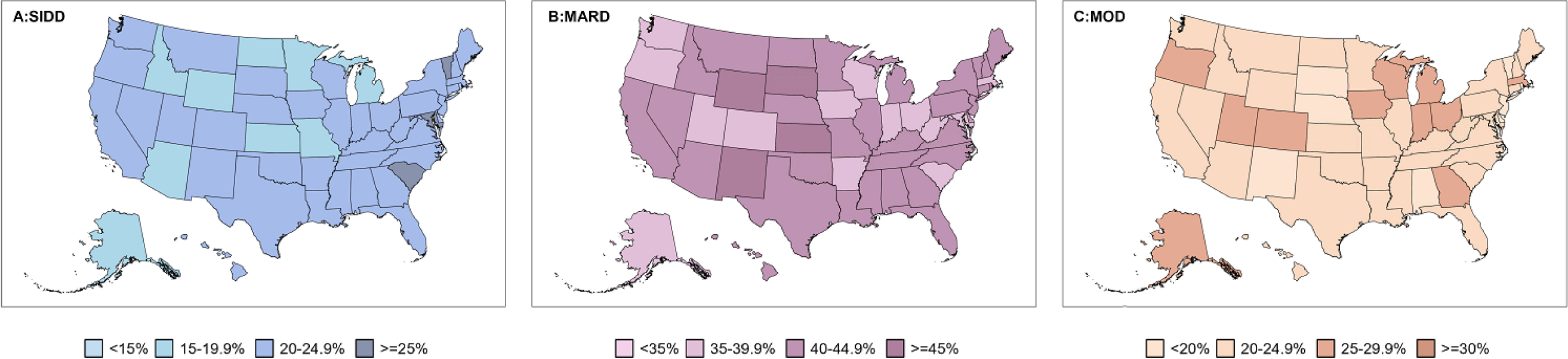
Geographic distribution of type 2 diabetes subphenotypes in Epic Cosmos. Estimates are based on a sample of 727,076 observations. Panel A: SIDD, Panel B: MARD, Panel C: MOD;

We also observed regional variations in the proportions of SIDD and MOD by race & ethnicity, although we did not conduct statistical tests for these differences. For example, a high proportion of non-Hispanic Black and Hispanic patients were classified as SIDD (≥20%) and MOD (≥30%) across several states (**Supplementary Figure 5, Panels A and C**). In contrast, the proportion of MARD was highest among non-Hispanic White patients (**Supplementary Figure 5, Panel B**).

### Time to pharmacological treatments

We utilized longitudinal EHRs and cumulative incidence functions to estimate the time to prescription of glucose-lowering medications (**Figure 4**). Hazard ratios (HR), adjusting for sex and age at diagnosis, were estimated using Cox proportional hazard models to study the time to prescription of insulin, metformin, and incretin mimetics (GLP-1 RA or GLP-1 RA/GIP) with subphenotype membership, relative to those classified as MOD (**Supplementary Table 14**). Relative to MOD in the first 60 months after diagnosis, prescription of insulin (adjusted HR: 1.65; 95% CI: 1.62, 1.67) and incretin mimetics (adjusted HR: 1.22; 95% CI: 1.20, 1.24) were earlier among SIDD. SIDD were also less likely to receive metformin (adjusted HR: 0.92, 95%CI: 0.91-0.94). Relative to MOD, initiation of prescription of insulin (adjusted HR: 0.87; 95% CI: 0.86, 0.89), metformin (adjusted HR: 0.81; 95% CI: 0.80, 0.83), and incretin mimetics (adjusted HR: 0.42; 95% CI: 0.41, 0.43) were later among MARD. Those unclassified were less likely to receive insulin (adjusted HR: 0.96, 95% CI: 0.94, 0.98) and incretin mimetics (adjusted HR: 0.84, 95% CI: 0.82, 0.85) but were more likely to receive metformin (adjusted HR: 1.08, 95% CI: 1.06, 1.10).

**Figure 4.**
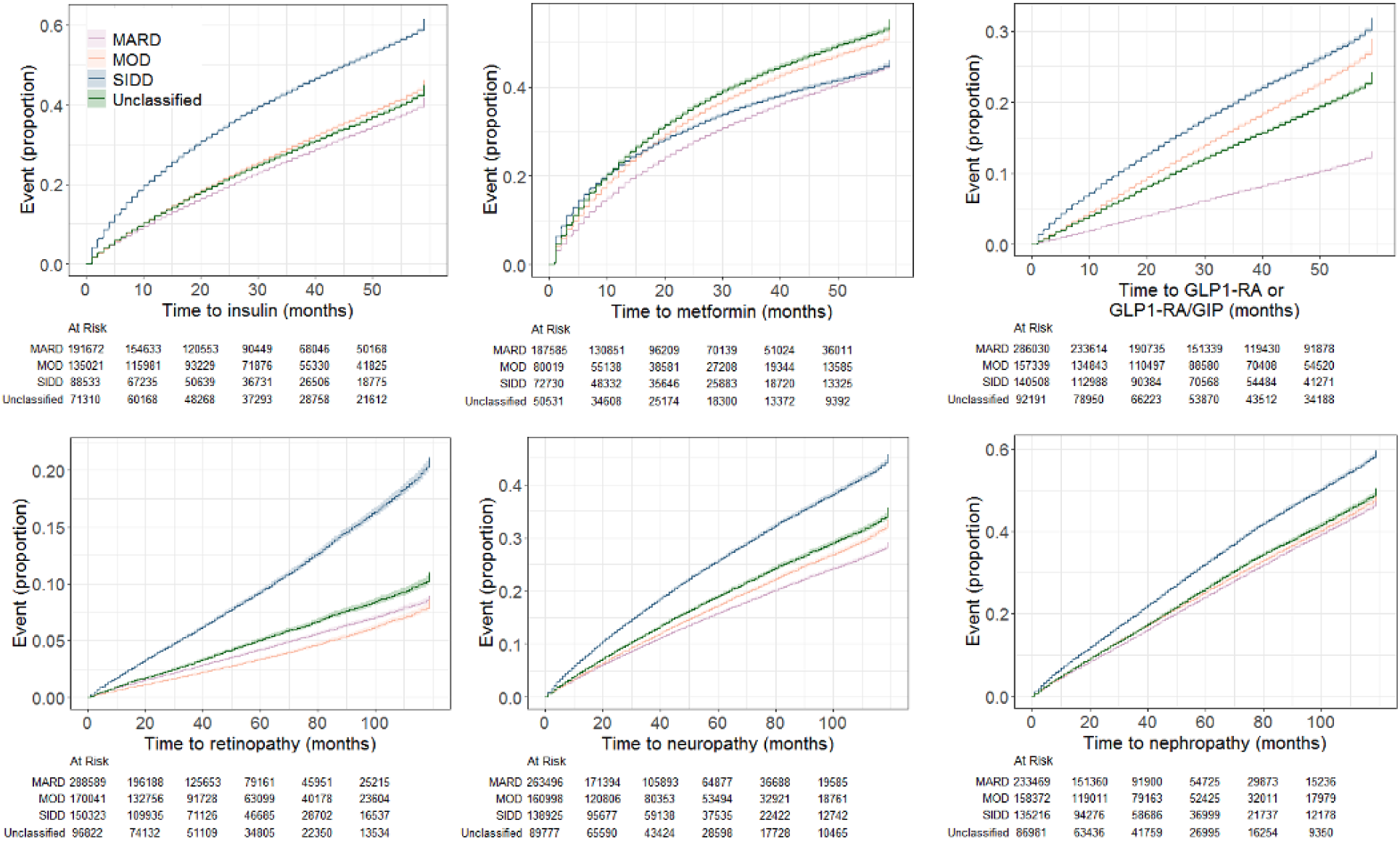
Time to pharmacological prescriptions and microvascular complications after diagnosis of type 2 diabetes in Epic Cosmos. All estimates are sex and age-adjusted cumulative incidence curves for (A) time to insulin, (B) time to metformin, (C) time to GLP1-RA or GLP1-RA/GIP (incretin mimetics), (D) time to retinopathy, (E)time to neuropathy and (F) time to nephropathy—more detailed information provided in **Supplementary Table 14.**

### Time to microvascular complications

We estimated the time to microvascular complications based on ICD-10-CM codes among patients free of these complication at diagnosis. SIDD exhibited a higher cumulative incidence of microvascular complications ten years after diagnosis **(Figure 4 and Supplementary Table 14)**. Relative to MOD in the first 10 years after diagnosis, SIDD were more likely to develop retinopathy (adjusted HR: 2.83, 95% CI: 2.73, 2.93), neuropathy (adjusted HR: 1.57, 95% CI: 1.54, 1.60) and nephropathy (adjusted HR: 1.34, 95% CI: 1.32, 1.37). Relative to MOD in the first 10 years after diagnosis, MARD were more likely to develop retinopathy (adjusted HR: 1.21, 95% CI: 1.16, 1.26) but less likely to develop neuropathy (adjusted HR: 0.90, 95% CI: 0.88, 0.92) and nephropathy (adjusted HR: 0.95, 95% CI: 0.93, 0.97). Those unclassified were more likely to develop retinopathy (adjusted HR: 1.43, 95% CI: 1.37, 1.49), neuropathy (adjusted HR: 1.10, 95% CI: 1.08, 1.13) and nephropathy (adjusted HR: 1.03, 95% CI: 1.01, 1.05). When comparing SIDD to all non-SIDD, the former were 2.41 times (95% CI: 2.36, 2.47), 1.61 times (95% CI: 1.59, 1.63) and 1.37 times (95% CI: 1.35, 1.38) more likely to develop retinopathy, neuropathy and nephropathy **(Supplementary Figure 6).**

## DISCUSSION

In this study, we successfully translated subphenotypes first identified in European cohort studies to a diverse EHR population from the US. By identifying four subphenotypes from newly diagnosed T2DM within pooled cohort studies from the United States, we developed and validated reliable classification models for detecting the novel subphenotypes of SIDD, MOD, and MARD. This replication and validation of diabetes subphenotypes in routine clinical care are critical steps towards successful translation of precision medicine into practice, since subphenotypes reflect variations in pathophysiology, treatment responses, and complication risks. The findings from this study have significant implications for diabetes surveillance and health policy.

Leveraging the largest integrated EHR database in the world, we revealed novel insights into the epidemiology of novel subphenotypes in the US. We estimated that one in five cases of newly diagnosed T2DM belong to the SIDD subphenotype, with higher proportions among racial and ethnic minorities. Estimates remain consistent with prior studies using smaller samples of new and previously diagnosed T2DM using national surveys and with those from a university health system.^17,19^ Furthermore, the prevalence of MARD and MOD subphenotypes in our study aligns with the patterns observed in other studies globally, where MARD is the most common subphenotype, particularly in non-Hispanic white populations, while MOD is more prevalent among younger individuals and those with higher body mass across racial and ethnic groups.^7^

Treatment patterns across subtypes revealed opportunities for optimizing diabetes care, prioritizing resource-limited treatments to high-risk groups. Notably, patients classified as SIDD were more likely to receive insulin and incretin mimetics (GLP-1 RA or GLP-1 RA/GIP) earlier than other subphenotypes during the follow-up, consistent with the ANDIS study and the American Diabetes Association’s recommendations for management of severe hyperglycemia.^20^ A recent randomized controlled trial demonstrated that stratifying patients into subtypes translated to tailored therapies that improved responses to semaglutide and dapagliflozin, with SIDD showing greater reductions in HbA1c and improved postprandial glucose control, highlighting the value of precision medicine in diabetes care ^21^. Although current approaches in clinical practice may prioritize those with insulin deficiency (SIDD), nearly half of SIDD cases were not treated with therapy appropriate to insulin deficiency within the first five years of diagnosis, indicating a gap in proper and timely treatment for this high-risk diabetic subgroup.

Conversely, overtreatment should be carefully managed, particularly among older adults in the mild age-related diabetes (MARD) subphenotype. Recent evidence suggests that overtreatment in older multimorbid patients can lead to adverse outcomes, such as higher mortality rates, without significant benefits in hospitalization or functional decline^22^. In our study, we observed that more than 20% of MARD received insulin treatment within the first two years of diagnosis. Alternatives such as GLP1-RAs may minimize the risk of falls from insulin-induced hypoglycemia in this population, provided concerns of loss of muscle mass and bone density are adequately studied.^23^ These results, therefore, underscore the need for research into a tailored treatment paradigm to address the unique T2DM management goals for each subphenotype.

We also observed differential risks of microvascular complications across subphenotypes, consistent with previous studies, highlighting the potential of precision prognosis for personalized microvascular complication prevention. SIDD exhibited a significantly higher incidence of diabetic retinopathy and neuropathy.^6,15^ However, the differences in cumulative incidence of nephropathy among SIRD, MOD and MARD were less distinguishable, relative to other studies.^6,24^ For instance, the risk of chronic kidney disease and albuminuria for SIDD was lower than SIRD and higher than MOD in the ANDIS study but was similar to MOD in the ADOPT trial.^6,24^

Findings from this study also have implications for diabetes surveillance and health policy. Current efforts towards achieving precision in public health skew towards genomics. Although useful for characterizing the burden and tailoring interventions for some forms of diabetes like monogenic diabetes of the young and potentially type 1 diabetes, genomics may be limited in its ability to stratify risk of complications after type 2 diabetes, a polygenic disease with only 19% of heritability explained by genetics.^27–29^ Most of the variability in treatment outcomes is, therefore, likely driven by structural socio-economic factors. For instance, we observed higher proportions of the SIDD subphenotype among non-Hispanic Black, Hispanic, and non-Hispanic Other adults, who also experience a higher prevalence of T2DM and worse social determinants of health. Geographic variability in SIDD proportions within racial and ethnic groups suggests the need for further investigation into the role of specific environmental factors and gene-environmental interactions contributing to the higher risk of SIDD in these populations. Additionally, individuals classified as SIDD and MOD reported higher social vulnerability, further complicating their diabetes management. Such insights could guide targeted interventions that address not only the biological underpinnings of diabetes but also the socio-economic factors influencing health outcomes.

This study has several strengths. First, we used data from rigorously conducted cohort studies with a low risk of undiagnosed diabetes or missing data due to left censoring of newly diagnosed T2DM. Second, we applied harmonized definitions across cohort studies, combining self-reported data and glycemic biomarkers to minimize information bias. Third, we utilized a validated computable phenotype of newly diagnosed T2DM and characterized the epidemiology and progression of a novel subphenotype, leveraging the largest integrated EHR database in the United States to characterize the epidemiology and progression of a novel subphenotype. Finally, our simple classification algorithm utilizing commonly available clinical measurements enables the clinical translation of our findings, facilitating practical application in routine clinical settings.

This study also has several limitations and challenges regarding subtyping diabetes for clinical validation. First, we excluded half of the newly diagnosed cases from the pooled cohort studies due to missing data, largely because of the absence of key biomarkers such as HbA1c in the earlier study visits in those cohort studies. While the analytic sample may not capture the full variability in newly diagnosed T2DM, the excluded and analytic samples were similar in distributions. Additionally, our methodological approach was designed to be robust against non-representativeness relative to the target population of all new T2DM cases. The data from six geographically diverse cohorts likely captures a broad phenotypical representation of diabetes cases. Moreover, applying our classification algorithm to a more representative large EHR dataset validated its robustness and enabled a nationally representative characterization of diabetes subphenotypes. Second, the SUPREME-DM computable phenotype does not specify the sequence of criterion (labs, medications, diagnostic codes) to meet for a case to be classified as incident T2DM in electronic health records. Therefore, nearly 1 in 3 cases and 1 in 6 cases had a history of insulin use and diabetic nephropathy, respectively, on or before the inclusion date in the analytic sample and were therefore excluded from the analysis of cumulative incidence. Third, the pooled cohorts might not fully represent the heterogeneity in diabetes within the US, particularly in its representation of other minorities such as Alaska Natives, Native Americans, South Asians and Pacific Islanders. Besides, although Epic Cosmos includes data from 250 million individuals in the US, only half the healthcare systems use Epic software.^30^ Nevertheless, a comparison of socio-demographic characteristics between Epic Cosmos and the US Census suggests similarities in socio-demographic characteristics.^31^ Finally, the unavailability of fasting insulin and fasting glucose in the EHRs prevented us from identifying SIRD. We hypothesize that distinguishing among the SIRD, MOD, and MARD subphenotypes may require additional biomarkers, such as liver and kidney function markers, to better capture the insulin-resistant pathophysiology.

Given the novel focus on integrating precision medicine into public health,^25^ we underscore two takeaways. First, prognostic models built using cohort studies and variables routinely collected in large electronic health record databases could enhance geographic surveillance of high-risk subphenotypes of newly diagnosed T2DM and monitor their prognosis.^26^ Second, the overlap of high-risk subphenotypes and social vulnerability emphasizes the importance of ensuring equitable access to high-quality diabetes care, including early diagnosis and access to highly efficacious medications like incretin mimetics, particularly in underserved communities, given their disproportionate burden of SIDD. This study, therefore, addresses critical gaps in translating T2DM subphenotypes into clinical practice, advancing opportunities for tailored treatment strategies. Our findings also support the development of predictive models for early subphenotype detection driving precision prevention. By enabling clinical validation, this research takes a key step toward integrating precision medicine into diabetes care.

## ONLINE METHODS

### Data Sources

#### Cohort studies

Data for subphenotyping of newly diagnosed T2DM consisted of six US-based longitudinal studies funded by the National Institutes of Health: Atherosclerosis Risk in Communities (ARIC), Coronary Artery Risk Development in Young Adults (CARDIA), Diabetes Prevention Program (DPP), DPP Outcomes Study (DPPOS is the long-term follow-up of DPP), Jackson Heart Study (JHS), and Multi-Ethnic Study of Atherosclerosis (MESA).^32–36^ Details of each cohort are provided in **Supplementary Note 1**. Our data, henceforth referred to as ‘pooled cohort studies’ consisted of a mix of observational cohorts (n =4) and follow-up of randomized trials (n=2).

#### Epic Cosmos

Data for validating the subphenotypes of newly diagnosed T2DM were from the Epic Cosmos Research Platform.^37^ Epic Cosmos includes HIPAA-compliant de-identified longitudinal electronic health records on nearly 250 million (at the time of analysis) unique patients from all 50 states of the United States, the District of Columbia, and Lebanon. Patients were linked longitudinally across health systems through an internal privacy-preserving process by Epic Cosmos. Socio-demographic variables were also harmonized across health systems by Epic Cosmos upon inclusion in the centralized platform and are broadly representative of the US population.^31^ Epic Cosmos additionally limited the geographic resolution to the state level and date-shifted the encounters at a patient level to prevent re-identification.

### Newly Diagnosed Type 2 Diabetes Mellitus

#### Pooled Cohort Studies

A newly diagnosed diabetes case definition was harmonized across the different cohorts based on the combination of questionnaire and biomarker data. Participants were classified as having newly diagnosed T2DM if they did not have a T2DM diagnosis at enrollment and either self-reported a new T2DM diagnosis, reported a diagnosis by a physician or health provider, or used diabetes medications in the last year (**Supplementary Table 3**). In ARIC, MESA, JHS, and CARDIA, participants who indicated diabetes at baseline but had missing age of diagnosis or diabetes duration or the earliest age of diagnosis was more than one year earlier than the baseline visit age were classified as having previously diagnosed diabetes. In the DPP trial and DPPOS, the development of diabetes was a primary outcome, so all diabetes cases were considered newly diagnosed. For those who did not self-report a diagnosis of diabetes, we relied on the 2024 ADA diagnostic criterion for identifying the cases, namely HbA1c ≥6.5% or fasting plasma glucose ≥126 mg/dL; or 2-hour oral glucose tolerance test ≥200 mg/dL (**Supplementary Table 1**). High random glucose was not considered as criteria for detecting new onset T2DM since data on diabetes symptoms were not available. We excluded all individuals for whom age, body mass index (BMI) and HbA1c were missing at diagnosis of T2DM (n = 3,390) or for whom homeostatic indices of beta cell function and insulin resistance were implausible (n = 13, acceptable input range for HOMA2 %B and HOMA2-IR calculation: insulin [20 to 400 pmol/L], glucose [3.0 to 25.0 mmol/L]). The analytic sample of newly diagnosed T2DM for the pooled cohort dataset consisted of 3,377 individuals (**Supplementary Figure 1**).

#### Epic Cosmos

Newly diagnosed type 2 diabetes was identified using the SUPREME-DM computable phenotype based on inpatient diagnosis codes or any combination of labs, outpatient diagnosis codes (International Classification of Diseases-10 or ICD-10-CM) and diabetes medications occurring within two years of each other (**Supplementary Table 13**).^38,39^ We considered the date of detection of the second criterion of SUPREME-DM as the date of diagnosis. To identify new-onset T2DM, we restricted our analysis to adult patients (18-99 years) who had in-person encounters (inpatient or outpatient) in each of the two years before detection. The final analytic sample of newly detected type 2 diabetes in Epic Cosmos consisted of 727,076 patients (**Supplementary Table 15**).

### Data Collection

#### Pooled Cohort Studies

We identified and obtained additional variables relevant for clinical phenotyping at or within one year after T2DM diagnosis. Nine clinical variables that are routinely measured during clinical visits were extracted, namely age at diagnosis, body mass index (BMI, kg/m²), systolic blood pressure (SBP), diastolic blood pressure (DBP), Hemoglobin A1c (HbA1c), Low-Density Lipoprotein (LDL) cholesterol, High-Density Lipoprotein (HDL) cholesterol, triglycerides, and the triglyceride-to-HDL cholesterol ratio (an indirect marker of insulin resistance).^18^ Measurement protocols and availability of key clinical variables for each cohort are presented in **Supplementary Table 2** and **Supplementary Table 4**. Additionally, demographic information such as gender, race and ethnicity, education level, and lifestyle factors, including drinking and smoking status, were harmonized across cohorts.

#### Epic Cosmos

Anthropometry (body mass index, blood pressure) and laboratory parameters (HbA1c, LDL cholesterol, HDL cholesterol, triglycerides) at the most recent visit in the year after detection of the SUPREME-DM computable phenotype were extracted. We additionally extracted other clinical and socio-demographic characteristics prior to the index visit: year of birth, biological sex (male, female), race-ethnicity (Hispanic, NH White, NH Black, NH Other), insurance status (Medicare, Medicaid, Private/unspecified, Self-pay) for the month of diagnosis, history of comorbidities based on ICD-10-CM codes, and prescriptions for the year prior to diagnosis. The percentile ranking for socio-economic position for the zip code of residence was available through linked Social Vulnerability Index 2020 data from the Center for Disease Control & Prevention. The most recent residential location was categorized as urban or rural based on US Department of Agriculture’s Rural-Urban Commuting Area 2010 primary codes.

### Data Cleaning

We harmonized variable names and units for the cardiometabolic biomarkers across the pooled cohorts: HbA1c was measured in percentages (%); blood pressure in millimeters of mercury (mmHg); LDL cholesterol, HDL cholesterol, triglycerides, and fasting blood glucose in milligrams per deciliter (mg/dL); and fasting insulin in micro-international units per milliliter (μIU/mL). Data from different studies were then integrated using common identifiers and harmonized definitions of variables. For the pooled cohort studies, we estimated the Homeostasis Model Assessment indicators, HOMA2 %B and HOMA2-IR, based on fasting insulin and fasting blood glucose, using the calculator published by the University of Oxford.^40^ For EHRs, vitals and laboratory parameters are harmonized by Epic Cosmos across all participating health systems. K-nearest neighbor imputation (k = 5) was used to impute missing values of cardiometabolic biomarkers in pooled cohorts and electronic health records.

### Classification and Statistical Analysis

#### Subphenotypes of Newly Diagnosed Type 2 Diabetes in Pooled Cohorts

We conducted an initial hierarchical clustering analysis using five variables (age of diagnosis, BMI, HbA1c, HOMA2 %B, and HOMA2 IR). Next, we conducted a k-means clustering analysis with varying numbers of clusters (k ranging from 2 to 10) and identified the optimal number using the Kneedle algorithm.^41^ The optimal number of clusters was identified as four from both unsupervised approaches. We used the clusters from the k-means clustering and labelled them based on their clinical similarity to the original ANDIS study subphenotypes as Mild Age-Related Diabetes (MARD), Mild Obesity-Related Diabetes (MOD), Severe Insulin-Deficient Diabetes (SIRD), and Severe Insulin-Resistant Diabetes (SIDD). The distribution of the five key variables is shown in **Supplementary Figure 3**.

#### Classification Model for Subphenotypes in Electronic Health Records

We constructed one-vs-all logistic regression models to predict the probability of being classified into each of the four subphenotype as a function of the nine routine clinical variables (age of diagnosis, BMI, HbA1c, SBP, DBP, LDL, HDL, triglycerides, triglyceride-to-HDL ratio). For example, the SIDD model estimated the probability of an individual being classified as SIDD and non-SIDD as a function of the nine variables. The pooled cohort data were split into 70% training and 30% test datasets. We identified the probability threshold for each model that maximized the F1 score (a combination of sensitivity and positive predictive value [PPV]). The models were used to predict subphenotype membership in the held-out test data. We evaluated the performance of our prediction models, relative to the cluster analysis labels, using the Area Under the Receiver Operating Characteristic Curves (AUC), sensitivity, specificity, PPV, negative predictive value (NPV) and F1 score. We applied the logistic regression models to the data from Epic Cosmos to sequentially estimate the probability of membership and classify cases into SIDD, MOD and MARD subphenotypes, given the low specificity of the SIRD model. The distribution of the nine clinical variables used in the logistical regression models is shown in **Supplementary Figure 4**.

#### Time to pharmacological Treatment and Risk of Microvascular Complications

In Epic Cosmos, we estimated cumulative incidence curves for prescription of glucose-lowering medication classes (insulin, metformin and incretin mimetics - glucagon-like peptide-1 receptor agonists or glucagon-like peptide-1 receptor agonists/glucose-dependent insulinotropic polypeptide). We also estimated the cumulative incidence of new-onset microvascular complications based on diagnostic codes (diabetic nephropathy [E11.2], retinopathy [E11.3], and neuropathy [E11.4]) among those free of each complication at diagnosis. Unadjusted Kaplan-Meier curves are shown in **Supplementary Figure 7 and 8**. We estimated hazard ratios and plotted survival curves of each of these outcomes for membership in SIDD, MARD, and unclassified cases after adjusting for sex, age, or both, relative to MOD (**Figure 4**). Because of the elevated risks associated with the SIDD subphenotype and its classification model’s high performance, we compared SIDD to all other subphenotypes grouped into non-SIDD, adjusting for age and sex (**Supplementary Figure 6**).

#### Sensitivity Analysis

First, to evaluate if one large and non-representative cohort influenced the final clustering, we assessed the concordance using the Adjusted Rand Index and Cohen’s κ between the original classification using the pooled cohort studies and those generated using a leave-one-cohort-out clustering approach. Second, we clustered the participants based on the nine routinely collected variables and compared these new clusters as an alternative to the original clusters.

Third, we trained multiclass prediction models to complement the clustering results as well as to assess whether non-linearity and statistical interactions between covariates can affect discrimination. We trained multinomial regression and random forest models using the training dataset. There were limited improvements in model performance across all subphenotypes (**Supplementary Table 13**). We observed that misdiagnosed SIRD cases were predominantly classified as MOD or MARD. A framework of the analysis plan is provided in **Supplementary Figure 1**. All analysis was conducted using Python 3.12.1 and R 4.2.3.

## Ethics approval and consent to participate

We were exempt from ethical approval for analysis of secondary datasets. All participants of cohort studies and trials gave written informed consent before participation. Epic Cosmos data was HIPAA-limited and expertly de-identified.

## Consent for publication

Not applicable

## Data availability

The code for the analysis is available on https://github.com/jvargh7/diabetes_endotypes_cohorts. Data for the National Institutes of Health funded cohort studies are available for request from the NIDDK Central Repository (NIDDK-CR; https://repository.niddk.nih.gov) and NHLBI Biolincc (https://biolincc.nhlbi.nih.gov/home/) to registered users. Epic Cosmos access is available through institutional representatives of participating institutions and the Epic Cosmos team after completing certification requirements.

## Author contributions

JSV and ZL conceptualized the study. JSV, ZL and SL developed the analytic plan with inputs from JCH. ZL and JSV led the data extraction, analysis and wrote the first draft. All authors reviewed and edited the subsequent drafts.

## Acknowledgments

We thank Kayla Yates, Meghan Howat, and Danessa Sandman of the Epic Cosmos team for their support.

