## Supplemental for "Translating Subphenotypes of Newly Diagnosed Type 2 Diabetes from Cohort Studies to Electronic Health Records in the United States"

**Supplementary Table 1. Description of National Institutes of Health-funded cohort studies**

| **Cohort** | **Study Years** | **Study Type** | **Description** | **Study website** |
| --- | --- | --- | --- | --- |
| Atherosclerosis Risk in Communities (ARIC) Study | 1985-Present | Cohort | The ARIC Study investigates the etiology and natural history of atherosclerosis and its clinical outcomes, including heart disease, stroke, and cognitive decline. The study involves approximately 16,000 participants aged 45-64 at baseline from four U.S. communities (Washington County, MD; Forsyth County, NC; Jackson, MS; and Minneapolis, MN). Participants undergo comprehensive examinations, including cardiovascular assessments, laboratory measurements, and lifestyle surveys. The study conducts follow-ups every three years to monitor changes in health status and risk factors. Notable findings include insights into the relationship between risk factors like hypertension, diabetes, cholesterol levels, and the incidence of cardiovascular diseases. | https://aric.cscc.unc.edu/aric9/ |
| Coronary Artery Risk Development in Young Adults Study (CARDIA) | 1985-Present | Cohort | The CARDIA Study focuses on understanding the development and determinants of cardiovascular disease in young adults. It enrolled over 5,000 black and white participants aged 18-30 at baseline, from four U.S. cities (Birmingham, AL; Chicago, IL; Minneapolis, MN; and Oakland, CA). The study includes extensive cardiovascular assessments, lifestyle and behavioral surveys, and psychosocial measures. Follow-up examinations occur every 2-5 years. Signific6ant findings have highlighted the impact of lifestyle factors such as diet, physical activity, and smoking on cardiovascular health, as well as the role of socioeconomic and psychosocial factors in disease risk. | https://www.cardia.dopm.uab.edu/ |
| Diabetes Prevention Program (DPP) | 1996-2001 | RCT | The DPP aimed to determine whether lifestyle intervention or the medication metformin could prevent or delay the onset of type 2 diabetes in high-risk individuals. The study included 3,234 participants with elevated fasting glucose or impaired glucose tolerance. Interventions focused on weight loss through diet and physical activity or metformin treatment. The study found that lifestyle intervention reduced diabetes incidence by 58%, and metformin by 31%, compared to placebo. Follow-ups assessed changes in diabetes status, metabolic measures, and cardiovascular risk factors. | https://www.niddk.nih.gov/about-niddk/research-areas/diabetes/diabetes-prevention-program-dpp |
| Diabetes Prevention Program Outcomes Study (DPPOS) | 2002 – present | RCT Follow-up of DPP | The DPPOS is a long-term follow-up of DPP participants to evaluate the sustained effects of the initial interventions on diabetes prevention and related health outcomes. It examines the long-term impact on diabetes incidence, cardiovascular outcomes, microvascular complications, and quality of life. The study continues to provide insights into the durability of lifestyle and pharmacologic interventions in managing and preventing type 2 diabetes over extended periods. | https://dppos.bsc.gwu.edu/ |
| Jackson Heart Study (JHS) | 1998-Present | Cohort | The JHS is a large single-site, prospective epidemiologic investigation focusing on identifying genetic and environmental risk factors associated with the disproportionate burden of cardiovascular disease in African Americans. The study involves around 5,300 participants from Jackson, Mississippi, metropolitan area of Hinds, Madison, and Rankin Counties. The initial baseline examination took place from 2000 to 2004, followed by two subsequent examinations from 2005 to 2008 and from 2009 to 2012. Follow-up examinations are conducted every three to four years, with ongoing annual follow-up interviews and cohort surveillance to track health outcomes and changes in risk factors. | https://www.jacksonheartstudy.org/ |
| Multi-Ethnic Study of Atherosclerosis (MESA) | 2000-Present | Cohort | MESA investigates the prevalence, causes, and progression of subclinical cardiovascular disease in a diverse, multi-ethnic population. The study includes over 6,800 participants from six U.S. communities (Los Angeles, CA; St. Paul, MN; New York, NY; Baltimore, MD; Chicago, IL; and Winston-Salem, NC), with no clinical cardiovascular disease at baseline. Assessments involve extensive imaging studies (e.g., CT scans, MRI) and biomarker measurements. Follow-up exams every two to four years monitor the development of cardiovascular disease and risk factors. The study has contributed to understanding ethnic differences in cardiovascular risk and the role of subclinical disease in predicting clinical events. | https://www.mesa-nhlbi.org/ |

**Supplementary Table 2. Measurement protocol for key biomarkers and anthropometry**

| **Cohort** | **Measure** | **Protocol** |
| --- | --- | --- |
| ARIC | Height and Weight | Standing height and weight are measured at enrollment and most study visits. Scale to measure body weight used in Detecto Model #437. Metal anthropometric rulers in centimeters (200cm) used to measure height. |
| CARDIA | Height and Weight | Height (anthropometric ruler in centimeters or Stadiometer) and weight (Detecto Scale Model#68965) were measured at baseline and all study follow-up visits |
| DPP | Height and Weight | Both height and weight were measured at the baseline and end of study. Weight was measured every 6 months. |
| DPPOS | Height and Weight | Weight was measured at 6-month intervals. Height was measured at the first annual visit and every 5-6 years. |
| JHS | Height and Weight | Weight was measured by a balance scale (Detector, model #437) and height was measured by traditional wall mounted tape at baseline and follow-up study visits. Tanita TBF 300A Body Composition Analyzer scale was used to assess the comparability of measurements. |
| MESA | Height and Weight | Weight was measured using a Detecto Platform Balance Scale and height was measured with stadiometer or height ruler at each study visit. |
| ARIC | HbA1c (%) | HbA1c was measured using high-performance liquid chromatography (HPLC) methods, standardized to the Diabetes Control and Complications Trial assay. Visit 5 was the first visit at which HbA1c was measured. |
| CARDIA | HbA1c (%) | HbA1c was measured at some study exams. |
| DPP | HbA1c (%) | HbA1c was measured at baseline, end of study, and 6 or 12-month intervals. |
| DPPOS | HbA1c (%) | HbA1c was measured at annual visits. |
| JHS | HbA1c (%) | HbA1c was measured using a National Glycohemoglobin Standardization Program (NGSP) certified assay at study baseline visit and follow up visits. |
| MESA | HbA1c (%) | Fasting lipid profile was measured at each study visit. HDL cholesterol was measured using the cholesterol oxidase method (Roche Diagnostics, Indianapolis, IN, USA). Triglyceride concentrations were measured using a glycerol-blanked enzymatic method LDL cholesterol was calculated using the Friedewald formula. |
| ARIC | Fasting glucose | The Central Clinical Chemistry Laboratory at the University of Minnesota processed the frozen blood serum samples collected from field centers. Participants were asked to fast prior to venipuncture. Fasting glucose was measured at enrollment and some study visits. |
| CARDIA | Fasting glucose | Fasting glucose was measured by hexokinase ultraviolet method at some study exams. |
| DPP | Fasting glucose | A blood sample (after 12 hours of fasting) was taken from the arm to measure fasting glucose levels every 6 months. |
| DPPOS | Fasting glucose | Fasting glucose was measured at both mid-year and annual visits. |
| JHS | Fasting glucose | Fasting glucose (8 hours of fasting) was measured by glucose oxidase method as duplicates in the baseline study visit and follow up visits. |
| MESA | Fasting glucose | Blood sample drawn at fasted state at all study visits. |
| ARIC | Fasting insulin | The Central Clinical Chemistry Laboratory at the University of Minnesota processed the frozen blood serum samples collected from field centers. Participants were asked to fast prior to venipuncture. Fasting insulin was measured at enrollment and some study visits. |
| CARDIA | Fasting insulin | Fasting insulin was measured by radioimmunoassay at some study exams. |
| DPP | Fasting insulin | Fasting insulin and fasting plasma proinsulin were collected at baseline, end of study, and every 12 months. |
| DPPOS | Fasting insulin | Fasting insulin was measured at mid-year visits. |
| JHS | Fasting insulin | Fasting plasma insulin were measured by total immunoreactive assay and standardized to serum levels at baseline study visits and follow up visits. |
| MESA | Fasting insulin | Blood sample drawn at fasted state at all study visits. |
| ARIC | Lipid panel | Lipid measurements were performed by the Central Lipid Laboratory. At baseline and multiple study visits, plasma cholesterol, triglycerides, HDL cholesterol and LDL cholesterol, plasma ApoA-I and apoB were collected. |
| CARDIA | Lipid panel | Lipid assays were conducted at each study exam including total cholesterol by trinder-type method and determined enzymatically on the Abbot Spectrum (using Hitachi 917 – R1 cholesterol reagent); HDL cholesterol by trinder-type method and determined enzymatically after dextran sulfate –magnesium precipitation on the Abbot Spectrum; and, triglycerides by ultraviolet method and determined enzymatically on the Abbot Spectrum (using Hitachi 917 – R1Buffer/4- Chloropheno/Enzymes). LDL cholesterol was calculated using the Friedewald equation. |
| DPP | Lipid panel | Fasting lipid profile (total cholesterol, total triglyceride, HDL-cholesterol and derived LDL-cholesterol) was measured at baseline, end of study, and at 6 or 12-month intervals. In cases of hypertriglyceridemia, beta quantification specifically measuring LDL-cholesterol is performed. |
| DPPOS | Lipid panel | Fasting lipid profile (total cholesterol, total triglyceride, HDL-cholesterol and derived LDL-cholesterol) was measured annually. In cases of hypertriglyceridemia, beta quantification specifically measuring LDL-cholesterol is performed. |
| JHS | Lipid panel | Routine plasma lipid tests (cholesterol, triglycerides, and HDL-cholesterol) were conducted at each study visit. |
| MESA | Lipid panel | Blood lipids (total cholesterol, HDL and LDL cholesterol, and triglycerides) and lipoproteins were measured at the Central Lipid Laboratory. |
| ARIC | Blood Pressure | The sitting arm blood pressure is measured three times at each clinic visit. It takes about 10-15 minutes to make three blood pressure measurements including the initial five minutes rest. |
| CARDIA | Blood Pressure | Seated BP is measured three times at each clinic visit. The seated BP reading is the average of the second and third systolic and diastolic BPs calculated by computer. |
| DPP | Blood Pressure | Blood pressure was measured in baseline, end of study, and in arm every 6 months and in ankle every 12 months. |
| DPPOS | Blood Pressure | Blood pressure was measured in arm at mid-year and annual visits and in ankle annually. |
| JHS | Blood Pressure | Sitting blood pressure were measured in a resting state, using 2 measurements with a random zero sphygmomanometer at each study visit. |
| MESA | Blood Pressure | Resting blood pressure was measured in the right arm  in the seated position by an automated oscillometric method (Dinamap) at all study exams. Three readings were taken; the second and third readings were averaged to obtain the blood pressure levels used in analyses. |

*Y20 protocol was used for CARDIA.

**Supplementary Table 3. Criterion for defining type 2 diabetes in National Institutes of Health cohorts and Epic Cosmos.**

| **Cohort** | **Criteria** |
| --- | --- |
| ARIC | **Cohort:** fasting blood glucose ≥126 mg/dl (7.0 mmol/L), non-fasting blood glucose ≥200 mg/dl (11.1 mmol/L), self-reported physician diagnosis of diabetes, or self-reported use of diabetes medications. |
| CARDIA | **Cohort:** fasting blood glucose ≥126 mg/dl (7.0 mmol/L), non-fasting blood glucose ≥200 mg/dl (11.1 mmol/L), self-reported physician diagnosis of diabetes, or self-reported use of diabetes medications. |
| DPP | **RCT:** fasting plasma glucose level ≥126 mg/dL (7.0 mmol/L), or 75-gram OGTT resulting in 2-hour plasma glucose ≥ 200 mg/dL (11.1 mmol/L). |
| DPPOS | **RCT:** fasting plasma glucose level ≥ 126 mg/dL [7.0 mmol/L] or 2-hour plasma glucose ≥ 200 mg/dL [11.1 mmol/L], after a 75-gram OGTT, and confirmed with a repeat test |
| JHS | **Cohort:** current use of insulin or oral antidiabetic agent, OR self-report of physician’s diagnosis or fasting glucose ≥ 126 mg/dl, or hemoglobin A1c ≥ 6.5% |
| MESA | **Cohort:** fasting blood glucose ≥126 mg/dl (7.0 mmol/L) OR participants self-reporting a physician-diagnosed history of diabetes and the use of insulin or oral hypoglycemic medications. |
| Epic Cosmos | **Electronic Health Records**: SUPREME-DM Computable Phenotype of newly diagnosed T2DM defined based on inpatient diagnosis codes or any combination of labs, outpatient diagnosis codes (International Classification of Diseases-10 or ICD-10-CM) and diabetes medications occurring within two years of each other.  **Diagnosis codes**  ICD-10-CM: E11  **Prescriptions**  *Confounding Classes*: Biguanide, Thiazolidinedione (PPAR G agonist), GLP-1 RA, GLP-1/GIP combination  *Other Classes*: Insulin, Biguanide + Sulfonylurea, DPP4 inhibitors, sulfonylurea, DPP4 inhibitors + Biguanide, Amylin analog, AGI, Dopamine receptor agonists, TZD+Sulfonylurea, SGLT2 inhibitors, DPP4 inhibitors + Statin, DPP4 inhibitor + TZD, SGLT2 inhibitor + Biguanide, Glucocorticoid blocker, Insulin + GLP-1RA, SGLT-2 inhibitor + DPP4 inhibitor + Biguanide  **Labs**  *HbA1c* ≥ *6.5%*: 4548-4, 41995-2, 55454-3, 71875-9, 549-2, 17856-6, 59261-6, 62388-4, 17855-8  *Fasting glucose* ≥ *126 mg/dL*:  1558-6, 76629-5, 1556-0, 77145-1, 35184-1,14771-0 |

**Supplementary Table 4. Descriptive characteristics of newly diagnosed type 2 diabetes in National Institutes of Health cohorts included in analytic sample**

|  | **ARIC** | **CARDIA** | **DPP** | **DPPOS** | **JHS** | **MESA** |
| --- | --- | --- | --- | --- | --- | --- |
|  | 1,062 | 228 | 285 | 1,013 | 245 | 544 |
| Age at diagnosis (SD) | 75.3 (5.1) | 48.8 (4.9) | 52 (9) | 57.7 (9.7) | 54.8 (10.8) | 67 (9.5) |
| Female % | 582(54.8%) | 129 (56.6%) | 184 (64.6%) | 676 (66.7%) | 156(63.7%) | 291(53.5%) |
| **Race^a^** |  |  |  |  |  |  |
| *White* | 766 (72.1%) | 67 (29.4%) | 151 (53%) | 557 (55%) | - | 144 (26.5%) |
| *Black* | 296 (27.9%) | 161 (70.6%) | 69 (24.2%) | 244 (24.1%) | 245 (100%) | 184 (33.8%) |
| *Other* | - | - | 65 (22.8%) | 212 (20.9%) | - | 216 (39.7%) |
| **Cluster** |  |  |  |  |  |  |
| MARD | 970 (91.3%) | 20 (8.8%) | 50 (17.5%) | 318 (31.4%) | 68 (27.8%) | 86 (15.8%) |
| MOD | 38 (3.6%) | 191 (83.8%) | 225 (78.9%) | 629 (62.1%) | 156 (63.7%) | 30 (5.5%) |
| SIDD | 44 (4.1%) | 16 (7%) | 5 (1.8%) | 4 (0.4%) | 19 (7.8%) | 20 (3.7%) |
| SIRD | 10 (0.9%) | 1 (0.4%) | 5 (1.8%) | 62 (6.1%) | 2 (0.8%) | 408 (75%) |
| **Key Biomarkers** |  |  |  |  |  |  |
| HbA1c (%) | 6.1 (5.8, 6.6) | 6.6 (6, 6.9) | 6.4 (6, 6.8) | 6.1 (5.7, 6.4) | 6.5 (6.2, 6.9) | 6.5 (6.1, 6.8) |
| Body mass index | 30.6 (5.9) | 35.3 (6.9) | 35.9 (7) | 35.1 (7.2) | 35.7 (7.8) | 31.1 (6) |
| HOMA2-B (%) | 71.2 (52.2, 94.9) | 102.2 (77.5, 133.5) | 118.5 (91.3, 154.7) | 129.9 (99.9, 172.1) | 100.7 (69.2, 137.4) | 273 (196, 353) |
| HOMA2-IR | 1.7 (1.1, 2.6) | 2.4 (1.6, 3.4) | 3.6 (2.5, 5.2) | 3.7 (2.4, 5.4) | 2.2 (1.3, 3.4) | 7.4 (5.2, 10.9) |
| Systolic BP (mmHg) | 129.6 (18) | 106.2 (27) | 126.3 (14.7) | 124.1 (14.4) | 128.7 (17.8) | 126.9 (19.6) |
| Diastolic BP (mmHg) | 66.8 (10.7) | 99.5 (27.2) | 79.4 (9.2) | 75.6 (9.3) | 78.6 (10.9) | 70.5 (10.9) |
| LDL cholesterol (mg/dL) | 97.3 (33.2) | 118.6 (37.9) | 119 (28.7) | 111.7 (32.5) | 121.5 (39.8) | 106.4 (33.3) |
| HDL cholesterol (mg/dL) | 48.5 (12.7) | 47.2 (13.6) | 42.1 (10.8) | 46.8 (12.1) | 49.5 (12.6) | 49.2 (13.9) |
| Triglycerides (mg/dL) | 141.2 (76.9) | 150.7 (118.9) | 175.3 (98.1) | 155.6 (98.1) | 128.8 (212.9) | 142.3 (87.3) |
| TGL:HDL ratio | 2.6 (1.8, 4) | 2.8 (1.8, 4.3) | 3.6 (2.5, 5.7) | 3 (2, 4.5) | 2.1 (1.5, 3.2) | 2.5 (1.7, 4.1) |

Values are mean (standard deviation) or median (25th percentile, 75th percentile) for continuous variables, and counts (percentages) for categorical variables. a Ethnicity information was not available for some cohorts and hence not reported.

**Supplementary Table 5. Comparison of clustering results for de-novo clustering with K means and apply ANIDS coordinates on analytic sample**

|  | ***ANDIS coordinates Clusters*** | | | | |
| --- | --- | --- | --- | --- | --- |
| ***De-novo clusters*** | **SIDD** | **SIRD** | **MOD** | **MARD** | **All** |
| SIDD | 105 | 1 | 2 | 0 | 108 |
| SIRD | 7 | 462 | 8 | 11 | 488 |
| MOD | 89 | 67 | 1029 | 84 | 1269 |
| MARD | 97 | 9 | 12 | 1394 | 1512 |
| All | 298 | 539 | 1051 | 1489 | 3377 |

**Supplementary Table 6. Stability and concordance using leave-one-cohort-out approach**

|  | N | Adjusted Rand Index | Cohen’s κ |
| --- | --- | --- | --- |
| **Overall** | ***3,377*** | - | 1.00 (Ref) |
| ARIC | -1,062 | 0.49 | 0.69 |
| CARDIA | -228 | 0.98 | 0.99 |
| DPP | -285 | 0.97 | 0.98 |
| DPPOS | -1013 | 0.96 | 0.98 |
| JHS | -245 | 0.97 | 0.98 |
| MESA | -544 | 0.63 | 0.68 |

**Supplementary Table 7. Comparison of analytic sample and complete case sample**

|  | **All newly diagnosed cases** | **Analytic Sample** | **Complete Case Sample** |
| --- | --- | --- | --- |
|  | 7,623 | 3,377 | 2,775 |
| Age at diagnosis (SD) | 61.6 (12.7) | 63.4 (12.4) | 63.1 (12.5) |
| Female % | 3,597 (56.5%) | 2,018 (59.8%) | 1,670 (60.2%) |
| **Race^a^** |  |  |  |
| *White* | 3316 (52.1%) | 1685 (49.9%) | 1500 (54.1%) |
| *Black* | 2364 (37.1%) | 1199 (35.5%) | 906 (32.6%) |
| *Other* | 686 (10.8%) | 493 (14.6%) | 369 (13.3%) |
| **Cohort** |  |  |  |
| ARIC | 4352 (57.1%) | 1062 (31.4%) | 918 (33.1%) |
| CARDIA | 623 (8.2%) | 228 (6.8%) | 224 (8.1%) |
| DPP | 291 (3.8%) | 285 (8.4%) | 269 (9.7%) |
| DPPOS | 1100 (14.4%) | 1013 (30%) | 987 (35.6%) |
| JHS | 268 (3.5%) | 245 (7.3%) | 129 (4.6%) |
| MESA | 989 (13%) | 544 (16.1%) | 248 (8.9%) |
| **Key Biomarkers** |  |  |  |
| HbA1c (%) | 6.2 (5.8, 6.7) | 6.3 (5.8, 6.7) | 6.2 (5.8, 6.6) |
| Body mass index (kg/m^2^) | 32 (6.7) | 33.2 (7) | 33.4 (7) |
| HOMA2-B (%) | 93.2 (63.4, 134.2) | 108 (73.3, 156.4) | 108.4 (73.3, 156.9) |
| HOMA2-IR | 2.4 (1.5, 4.1) | 2.8 (1.7, 4.7) | 2.8 (1.7, 4.7) |
| Systolic BP (mmHg) | 135.8 (23.3) | 125.6 (18.6) | 124.9 (18.3) |
| Diastolic BP (mmHg) | 82 (17.5) | 74.2 (14.6) | 74.6 (15) |
| LDL cholesterol (mg/dL) | 116.8 (37.9) | 108 (34.5) | 107.7 (34.4) |
| HDL cholesterol (mg/dL) | 43.4 (16.6) | 47.6 (12.8) | 47.4 (12.7) |
| Triglycerides (mg/dL) | 159.2 (122.6) | 148.3 (104.3) | 145.7 (82.5) |
| TGL:HDL ratio | 3.1 (2, 5.4) | 2.8 (1.8, 4.2) | 2.8 (1.8, 4.2) |

Values are mean (standard deviation) or median (25th percentile, 75th percentile) for continuous variables, and counts (percentages) for categorical variables. Ethnicity information was not available for some cohorts and hence not reported.

**Supplementary Table 8. Comparison of clustering results for analytic sample and de-novo clustering of complete cases**

|  | ***Complete case analysis*** | | | | |
| --- | --- | --- | --- | --- | --- |
| ***Analytic Sample*** | **SIDD** | **SIRD** | **MOD** | **MARD** | **All** |
| SIDD | 77 | 0 | 0 | 0 | 77 |
| SIRD | 0 | 242 | 0 | 0 | 242 |
| MOD | 3 | 30 | 1086 | 16 | 1135 |
| MARD | 1 | 11 | 27 | 1282 | 1321 |
| All | 81 | 283 | 1113 | 1298 | 2775 |

**Supplementary Table 9. Missing data for analytic sample before k-nearest neighbor imputation**

| **Data Source** | **N** | **Age** | **BMI** | **HbA1c** | **Fasting glucose** | **Fasting Insulin** | **LDL** | **HDL** | **TGL** | **SBP** | **DBP** |
| --- | --- | --- | --- | --- | --- | --- | --- | --- | --- | --- | --- |
| ARIC | 1,062 | 0 | 0 | 0 | 69 | 112 | 13 | 0 | 0 | 4 | 4 |
| CARDIA | 228 | 0 | 0 | 0 | 0 | 0 | 4 | 0 | 0 | 0 | 0 |
| DPP | 285 | 0 | 0 | 0 | 0 | 9 | 7 | 7 | 7 | 0 | 0 |
| DPPOS | 1014 | 0 | 0 | 0 | 0 | 10 | 21 | 21 | 21 | 0 | 0 |
| JHS | 245 | 0 | 0 | 0 | 26 | 114 | 29 | 26 | 26 | 2 | 2 |
| MESA | 556 | 0 | 0 | 0 | 0 | 293 | 10 | 0 | 0 | 0 | 0 |
| Epic Cosmos | 727,076 | 0 | 70,145 | 73,443 | 706,688 | 726,756 | 231,333 | 246,713 | 240,568 | 24,859 | 24,859 |

Values are counts of missing observations for each biomarker

**Supplementary Table 10. Comparison of cluster results for five variable and nine variable methods for four clusters**

|  | **Cluster 1** | **Cluster 2** | **Cluster 3** | **Cluster 4** | **All** |
| --- | --- | --- | --- | --- | --- |
| SIDD | 17 | 29 | 9 | 53 | 108 |
| SIRD | 48 | 203 | 123 | 114 | 488 |
| MOD | 79 | 92 | 102 | 996 | 1269 |
| MARD | 56 | 769 | 623 | 64 | 1512 |
| All | 200 | 1093 | 857 | 1227 | 3377 |

**Supplementary Table 11. Coefficients and standard errors of one-versus-all logistic regression classification models**

|  | **SIDD** | **SIRD** | **MOD** | **MARD** |
| --- | --- | --- | --- | --- |
| Intercept | -51.89 (8.68) | -1.92 (0.90) | 8.64 (1.18) | -0.72 (1.05) |
| Age at diagnosis | 0.00 (0.04) | 0.01 (0.01) | -0.22 (0.01) | 0.18 (0.01) |
| HbA1c (%) | 5.90 (0.93) | 0.04 (0.06) | -0.79 (0.09) | -0.86 (0.09) |
| Body mass index | 0.05 (0.05) | -0.01 (0.01) | 0.24 (0.02) | -0.18 (0.01) |
| Systolic BP | 0.03 (0.02) | 0.01 (0.00) | 0.01 (0.00) | 0.00 (0.00) |
| Diastolic BP | -0.08 (0.04) | -0.02 (0.01) | 0.01 (0.01) | 0.01 (0.01) |
| LDL cholesterol | 0.02 (0.01) | 0.00 (0.00) | 0.00 (0.00) | 0.00 (0.00) |
| HDL cholesterol | 0.04 (0.04) | 0.00 (0.01) | -0.02 (0.01) | 0.00 (0.01) |
| Triglycerides | -0.02 (0.01) | 0.00 (0.00) | 0.01 (0.00) | 0.00 (0.00) |
| TGL:HDL ratio | 0.44 (0.38) | 0.08 (0.06) | -0.21 (0.09) | -0.18 (0.09) |

All values are coefficients and standard errors (in parentheses) in logit scale.

**Supplementary Table 12. Evaluation of discrimination of type 2 diabetes subphenotypes using usual clinical variables in pooled cohort studies**

|  | **Sensitivity** | **Specificity** | **AUC** | **PPV** | **NPV** | **F_1_ Score** | **Probability Cutoff** ^a^ |
| --- | --- | --- | --- | --- | --- | --- | --- |
| **Training** |  |  |  |  |  |  |  |
| SIDD | 0.97 | 1.00 | 1.00 | 0.87 | 1.00 | 0.92 | 0.16 |
| SIRD | 0.83 | 0.31 | 0.58 | 0.17 | 0.92 | 0.28 | 0.12 |
| MOD | 0.92 | 0.87 | 0.95 | 0.81 | 0.95 | 0.86 | 0.38 |
| MARD | 0.95 | 0.81 | 0.93 | 0.80 | 0.95 | 0.87 | 0.32 |
| **Test^b^** |  |  |  |  |  |  |  |
| SIDD | 0.94 | 0.99 | 1.00 (0.99, 1.00) | 0.86 | 1.00 | 0.90 | - |
| SIRD | 0.85 | 0.30 | 0.62 (0.58, 0.67) | 0.17 | 0.92 | 0.28 | - |
| MOD | 0.93 | 0.89 | 0.96 (0.95, 0,.97) | 0.84 | 0.96 | 0.88 | - |
| MARD | 0.95 | 0.81 | 0.93 (0.92, 0.95) | 0.80 | 0.95 | 0.87 | - |

All estimates are cutoff maximizing F1 score in the training dataset. Models were fit first on a training dataset (n = 2,363 ) and performance was assessed on a held-out test dataset (n = 1,014 ). AUC: Area under receiver operating characteristic curve; PPV: Positive Predictive Value; NPV: Negative Predictive Value.

a Probability cutoff based on maximizing F_1_ score in the training dataset.

b The 95% confidence interval is derived from the 2.5th and 97.5th percentiles of the AUC distribution from resampling the test data with replacement (bootstrap n = 1000).

**Supplementary Table 13. Performance of supervised classification models in training data**

|  | **Sensitivity** | **Specificity** | **AUC** | **PPV** | **NPV** | **F1-Score** |
| --- | --- | --- | --- | --- | --- | --- |
| ***Multinomial*** |  |  |  |  |  |  |
| SIDD | 0.84 | 1.00 | 1.00 | 0.91 | 0.99 | 0.88 |
| SIRD | 0.09 | 0.97 | 0.69 | 0.30 | 0.86 | 0.13 |
| MOD | 0.94 | 0.90 | 0.97 | 0.85 | 0.96 | 0.90 |
| MARD | 0.94 | 0.83 | 0.94 | 0.81 | 0.94 | 0.87 |
| ***Random Forests*** |  |  |  |  |  |  |
| SIDD | 0.96 | 1.00 | 1.00 | 0.91 | 1.00 | 0.94 |
| SIRD | 0.10 | 0.98 | 0.71 | 0.47 | 0.87 | 0.16 |
| MOD | 0.95 | 0.90 | 0.97 | 0.85 | 0.97 | 0.90 |
| MARD | 0.94 | 0.82 | 0.94 | 0.81 | 0.95 | 0.87 |

**Supplementary Table 14. Time to prescription and microvascular complications among newly diagnosed type 2 diabetes cases in Epic Cosmos**

|  |  | **Excluded** | **Events (n)** | **Censored (n)** | **Follow-up (months)** | **Sex-adjusted HR (95% CI)** | **Sex and age-adjusted HR (95% CI)** |
| --- | --- | --- | --- | --- | --- | --- | --- |
| **Insulin** | **MOD** | 592,055 | 35,917 | 99,104 | 32.9 | 1 (ref) | 1 (ref) |
|  | **MARD** | 535,404 | 61,384 | 130,288 | 28.0 | 1.32 (1.3, 1.33) | 0.87 (0.86, 0.89) |
|  | **SIDD** | 638,543 | 36,958 | 51,575 | 24.9 | 1.88 (1.85, 1.91) | 1.65 (1.62, 1.67) |
|  | **Unclassified** | 655,766 | 21,835 | 49,475 | 31.9 | 1.17 (1.15, 1.19) | 0.96 (0.94, 0.98) |
| **Metformin** | **MOD** | 647,057 | 35,541 | 44,478 | 19.0 | 1 (ref) | 1 (ref) |
|  | **MARD** | 539,491 | 45,826 | 141,759 | 21.0 | 0.53 (0.52, 0.54) | 0.81 (0.80, 0.83) |
|  | **SIDD** | 655,346 | 24,774 | 47,956 | 19.9 | 0.77 (0.76, 0.78) | 0.92 (0.91, 0.94) |
|  | **Unclassified** | 676,545 | 20,042 | 30,489 | 20.0 | 0.87 (0.86, 0.89) | 1.08 (1.06, 1.1) |
| **Incretin Mimetics** | **MOD** | 569,737 | 45,203 | 112,136 | 35.9 | 1 (ref) | 1 (ref) |
|  | **MARD** | 441,046 | 18,805 | 267,225 | 32.9 | 0.24 (0.24, 0.25) | 0.42 (0.41, 0.43) |
|  | **SIDD** | 586,568 | 37,285 | 103,223 | 30.9 | 1.03 (1.02, 1.05) | 1.22 (1.2, 1.24) |
|  | **Unclassified** | 634,885 | 16,883 | 75,308 | 37.9 | 0.63 (0.62, 0.64) | 0.84 (0.82, 0.85) |
| **Retinopathy** | **MOD** | 557,035 | 4,899 | 165,142 | 44.0 | 1 (ref) | 1 (ref) |
|  | **MARD** | 438,487 | 10,058 | 278,521 | 33.9 | 1.49 (1.44, 1.55) | 1.21 (1.16, 1.26) |
|  | **SIDD** | 576,753 | 11,695 | 138,628 | 37.9 | 3.03 (2.93, 3.13) | 2.83 (2.73, 2.93) |
|  | **Unclassified** | 630,254 | 4,340 | 92,482 | 42.9 | 1.59 (1.53, 1.66) | 1.43 (1.37, 1.49) |
| **Neuropathy** | **MOD** | 556,078 | 19.492 | 141,506 | 39.9 | 1 (ref) | 1 (ref) |
|  | **MARD** | 463,580 | 35,675 | 227,821 | 31.0 | 1.35 (1.32, 1.37) | 0.9 (0.88, 0.92) |
|  | **SIDD** | 588,151 | 27,035 | 111,890 | 33.9 | 1.81 (1.77, 1.84) | 1.57 (1.54, 1.60) |
|  | **Unclassified** | 637,299 | 14,281 | 75,496 | 38.9 | 1.35 (1.32, 1.38) | 1.10 (1.08, 1.13) |
| **Nephropathy** | **MOD** | 568,704 | 23,759 | 134,613 | 40.0 | 1 (ref) | 1 (ref) |
|  | **MARD** | 493,607 | 60,970 | 172,499 | 30.9 | 2.16 (2.13, 2.20) | 0.95 (0.93, 0.97) |
|  | **SIDD** | 591,860 | 32,664 | 102,552 | 34.9 | 1.8 (1.77, 1.83) | 1.34 (1.32, 1.37) |
|  | **Unclassified** | 640,095 | 19,368 | 67,613 | 38.9 | 1.53 (1.50 1.56) | 1.03 (1.01, 1.05) |

**Supplementary Table 15. Descriptive characteristics of newly detected type 2 diabetes by subphenotype in Epic Cosmos**

|  |  | **Overall** | **Non-SIDD** | **SIDD** |
| --- | --- | --- | --- | --- |
|  | **Available N** | *727,076* | *570,125* | *156,951* |
| Age at detection of SUPREME-DM | 727,076 | 64.4 (13.3) | 65.6 (13.0) | 59.9 (13.5) |
| Female | 727,076 | 52% | 52% | 50% |
| **Race & Ethnicity** | 727,076 |  |  |  |
| *NH White* |  | 68% | 69% | 63% |
| *NH Black* |  | 17% | 16% | 20% |
| *Hispanic* |  | 7% | 6.4% | 9.3% |
| *NH Other* |  | 8.6% | 8.7% | 8.0% |
| Social Vulnerability Index (0: Low, 100: High vulnerability) | 721,964 | 60.4 (33.5, 82.1) | 59.4 (32.7, 81.3) | 64.1 (37.0, 84.6) |
| **Insurance** | 727,076 |  |  |  |
| *Medicare* |  | 40% | 42% | 31% |
| *Medicaid* |  | 14% | 13% | 16% |
| **Key Biomarkers** |  |  |  |  |
| HbA1c (%) | 653,633 | 7.0 (6.6, 7.9) | 6.8 (6.5, 7.2) | 9.5 (8.7, 11.0) |
| Body mass index | 656,931 | 33.2 (6.8) | 33.0 (6.8) | 34.0 (6.9) |
| Systolic BP | 702,217 | 131.2 (15.5) | 130.8 (15.3) | 132.5 (16.3) |
| Diastolic BP | 702,217 | 74.7 (10.2) | 74.3 (10.1) | 76.2 (10.4) |
| LDL cholesterol | 495,743 | 89.0 (37.8) | 87.0 (36.4) | 96.2 (41.7) |
| HDL cholesterol | 480,363 | 44.9 (14.6) | 45.5 (14.4) | 42.9 (15.1) |
| Triglycerides | 486,508 | 165.0 (92.4) | 159.3 (87.5) | 185.9(106.2) |
| TGL:HDL ratio | 467,823 | 3.3 (2.1, 5.3) | 3.2 (2.0, 5.0) | 3.9 (2.4, 6.4) |

Values are mean (standard deviation) or median (25^th^ percentile, 75^th^ percentile) for continuous variables, and percentages for categorical variables.

**Supplementary Figure 1. Analytic sample flowchart**

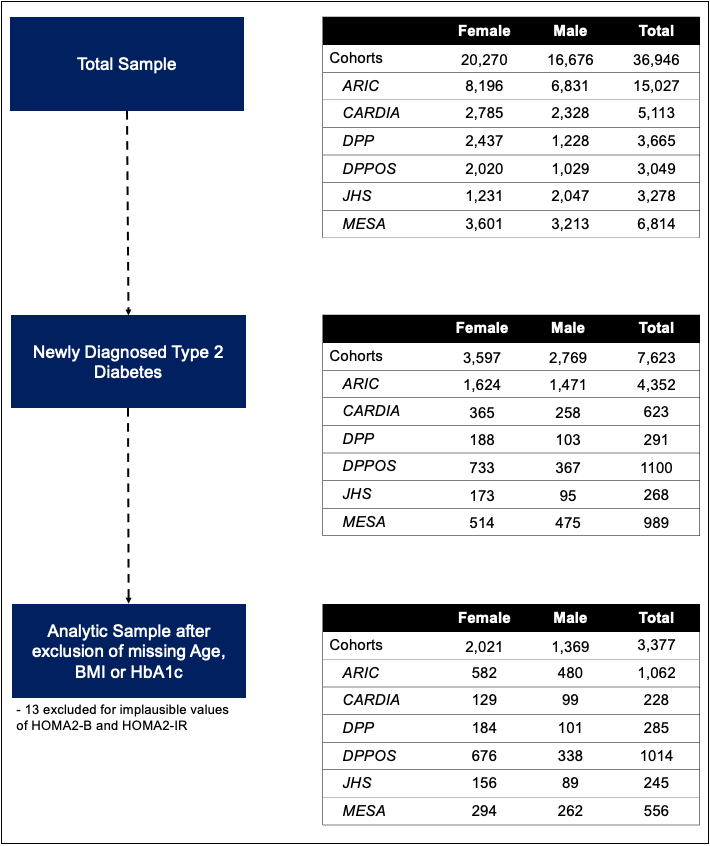

**Supplementary Figure 2. K-means clustering elbow plot**

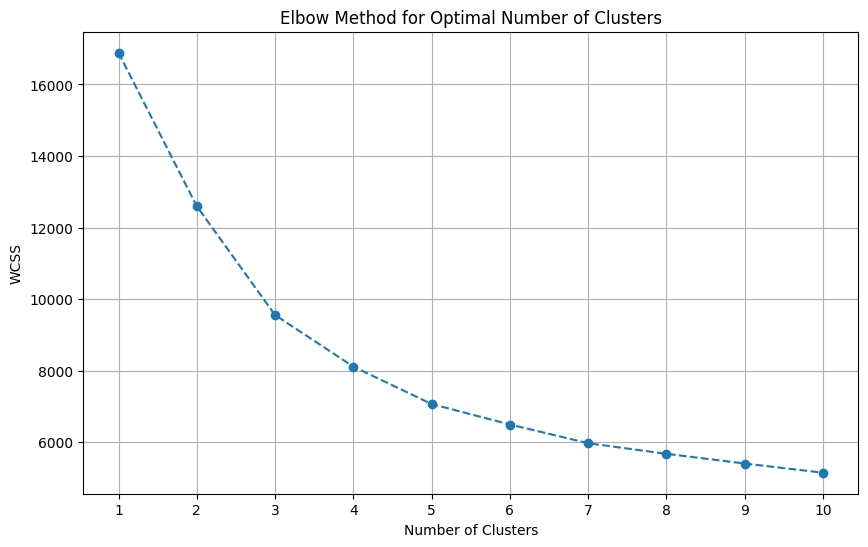

This elbow plot shows the within-cluster sum of squares (WCSS) against the number of clusters (k) for k-means clustering applied to five selected variables (BMI, HbA1c, age at diabetes diagnosis, HOMA2-B, and HOMA2-IR) in the de-novo classification of T2DM subphenotypes.

**Supplementary Figure 3. Distribution of variables in pooled cohorts by cluster after k-nearest neighbor imputation for prediction, n = 3,377**

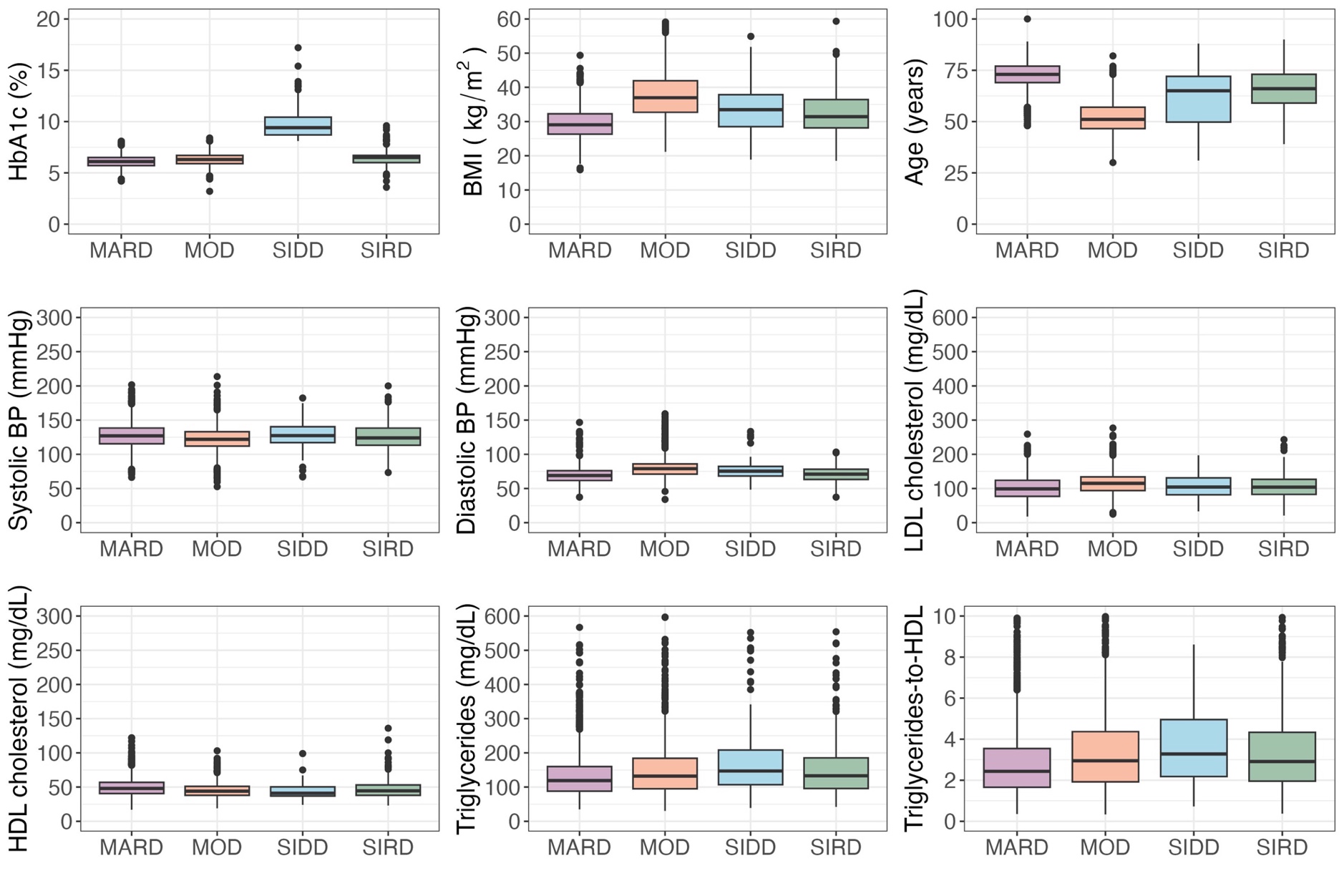

**Supplementary Figure 4. Distribution of variables in Epic Cosmos by subphenotypes of type 2 diabetes, n = 727,076**

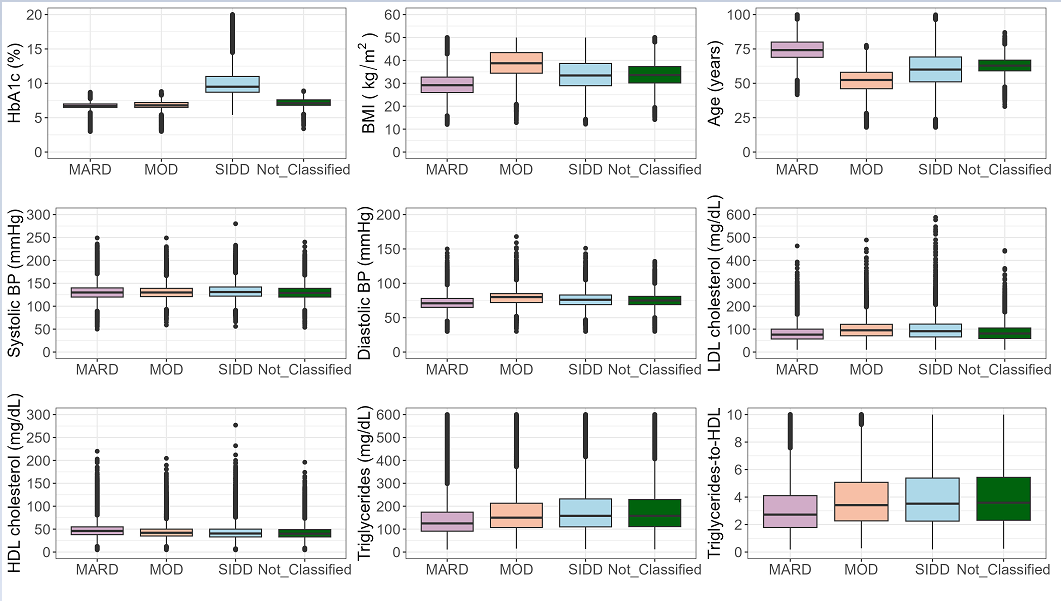

**Supplementary Figure 5. Proportion of subphenotypes of diabetes among newly diagnosed type 2 diabetes cases in Epic Cosmos**

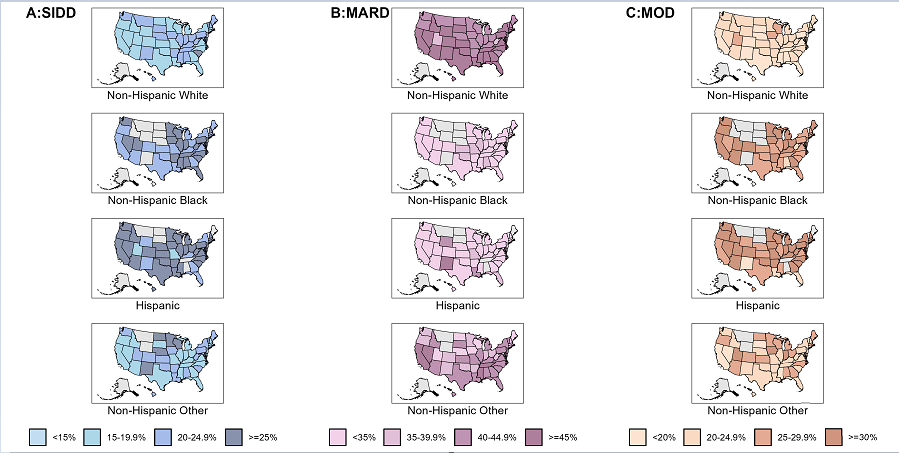

We used a rule-based approach to classify cases into SIDD, MOD and MARD, based on decreasing F_1_ scores on the test dataset (**Table 2**). Cases were first classified as SIDD if their probability of being identified as SIDD exceeded 0.16. Next, the remaining cases were classified into MARD if their probability of being identified as MARD exceeded 0.32. Finally, the remaining cases were classified as MOD if their probability of being identified as MOD exceeded 0.38.

Panel A: SIDD, Panel B: MARD, Panel C: MOD; Rows: Among Non-Hispanic White cases; Among Non-Hispanic Black cases; Among Hispanic cases; Among Non-Hispanic Other cases.

**Supplementary Figure 6. Time to pharmacological prescriptions and microvascular complications after diagnosis of severe insulin-deficient diabetes in Epic Cosmos**

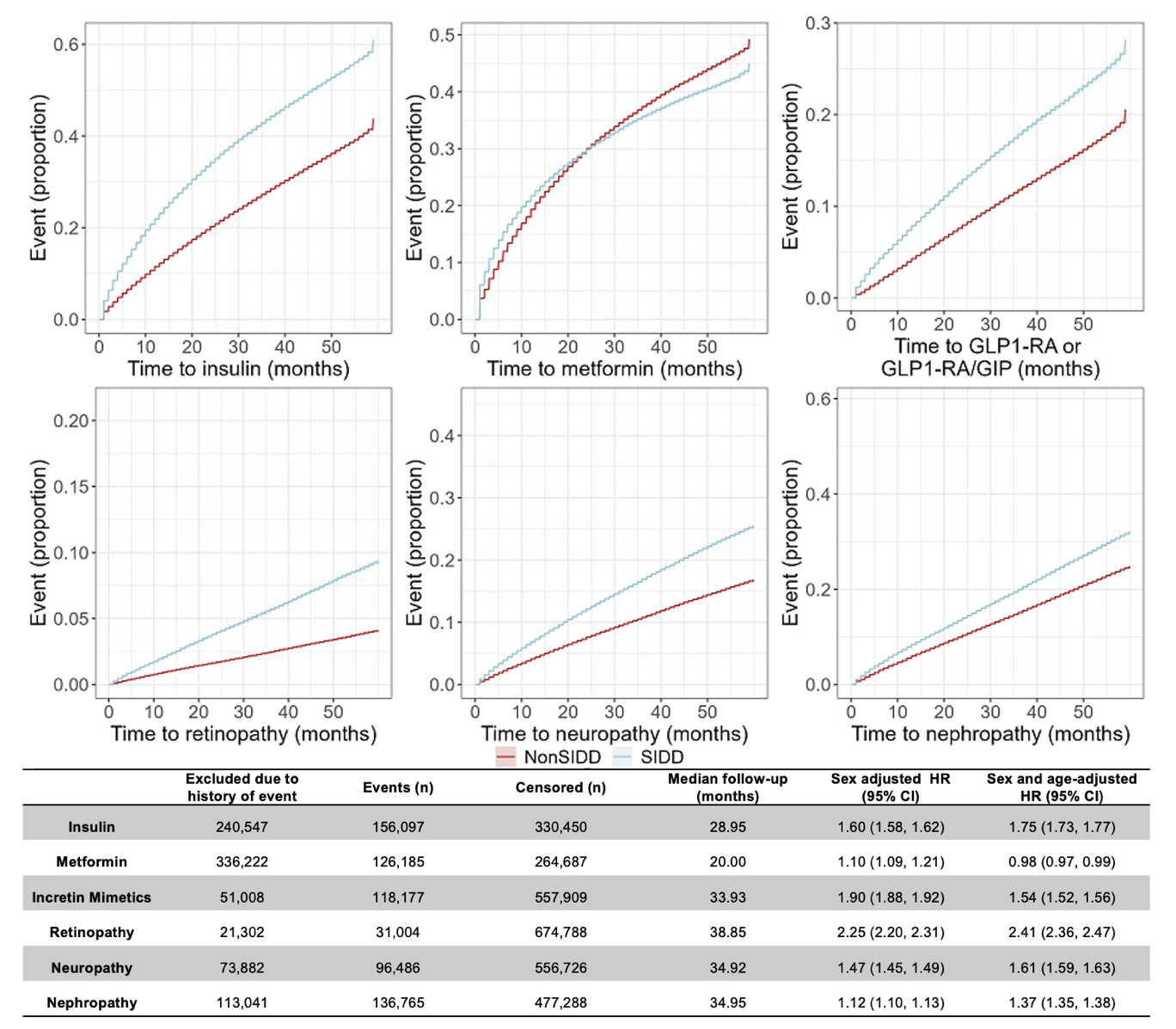

All estimates are unadjusted cumulative incidence curves for time to top panel: insulin, metformin, GLP1-RA or GLP1-RA/GIP (incretin mimetics); bottom panel: retinopathy, neuropathy and nephropathy.

**Supplementary Figure 7. Unadjusted Kaplan Meier Curves of Subphenotypes**

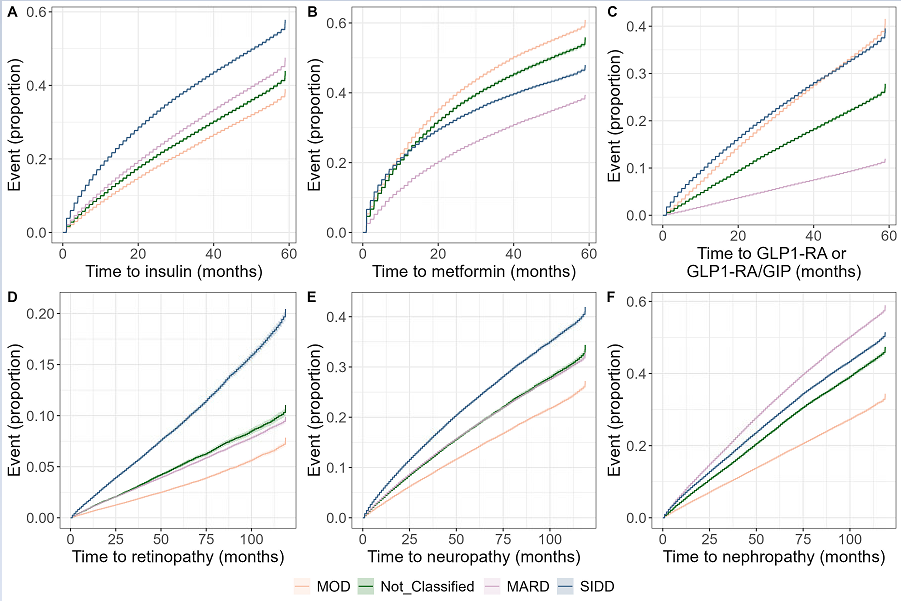

**Supplementary Figure 8. Unadjusted Kaplan Meier Curves of SIDD and non-SIDD Subphenotypes**

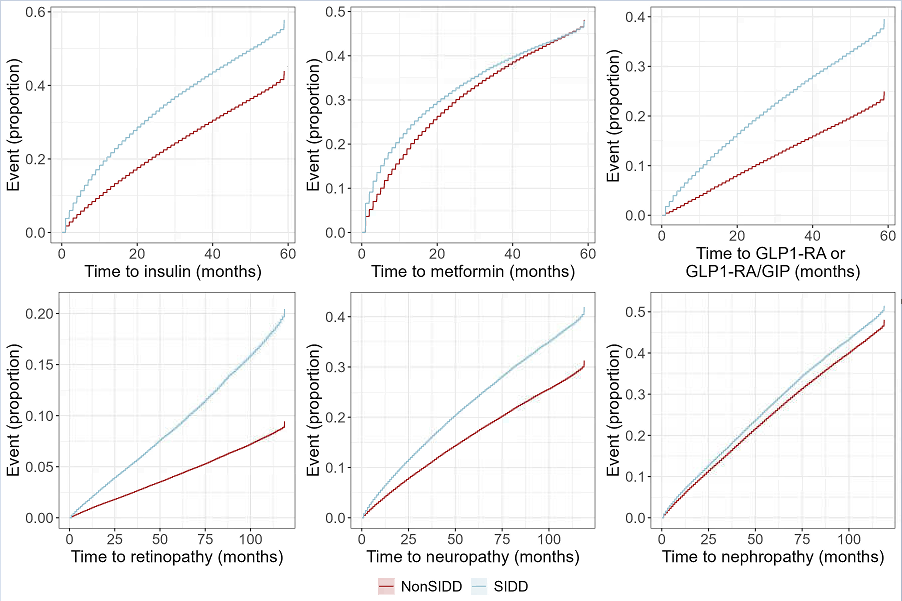
